# Effect of ancestry and shared genetic architecture of serious mental illness on symptoms and cognition in an admixed Latin American population

**DOI:** 10.64898/2026.05.05.26351986

**Authors:** Esteban Lopera-Maya, Susan K Service, Ana M. Díaz-Zuluaga, Mauricio Castaño Ramírez, Juan Camilo Mejia, Johanna Valencia, Terri Teshiba, Ana Maria Ramirez, Juan F De la Hoz, Jonathan Valdez, Marfred Munoz Umanes, Tyler M. Moore, Sinéad Chapman, Benjamin M. Neale, Carrie E. Bearden, Javier I. Escobar, Ruben C. Gur, Victor I. Reus, Chiara Sabatti, Loes Olde Loohuis, Carlos Lopez Jaramillo, Nelson B. Freimer

## Abstract

Most genome-wide association studies (GWAS) of serious mental illness (SMI) have been conducted for categorical diagnoses in samples of primarily European ancestry. The portability of findings to non-Europeans, and to SMI-related symptoms/dimensional traits remains uncertain. In a sample of 8,666 SMI cases and controls from the Paisa region of Colombia we show that a primarily European schizophrenia GWAS polygenic risk score (PRS) predicted all SMI diagnoses in this sample, as well as symptoms (assessed in cases only) and traits assessed agnostic to SMI diagnosis: a one SD unit (SDU) increase in this PRS was associated to decreased risk in cases of suicidal thoughts (OR=0.89, 95% confidence interval 0.84-0.94), depressed mood (OR=0.90, 95% confidence interval 0.85-0.95), and increased risk of delusions (OR=1.12, 95% confidence interval 1.06-1.18) and to decreased cognition (in cases and controls) across five distinct domains (average decrease in cognition of 0.065 SDU, p<7e-05). We show that a published European GWAS of cognition predicted levels of executive function (average decrease in cognition of 0.06 SDU per unit increase in PRS, p<2e-04), but not diagnosis or symptoms. Specific loci identified in the SMI GWAS also showed association to multiple diagnoses, symptoms, and cognitive traits in Paisa. The most noteworthy result was for a locus on chromosome 7p22.3, associated in multiple SMI GWAS, that showed association in Paisa to increased risk of bipolar disorder, and to reduced complex cognition and social cognition. Our findings demonstrate wide portability from European GWAS to an admixed American sample, with associations to multiple transdiagnostic phenotypes.

## Introduction

Genome wide association studies (GWAS) have identified hundreds of loci contributing to risk for serious mental illness (SMI) diagnoses, defined here as including schizophrenia (SCZ), bipolar disorder (BD), and major depressive disorder (MDD). These studies mostly include participants with predominantly European ancestry; it remains unclear how well their results translate to populations with other ancestries. Additionally, most SMI GWAS have utilized cases recruited separately for each diagnosis and have treated diagnoses as distinct lifetime categories. Several studies, however, have revealed substantial cross-disorder genetic correlations [1–3], identified transdiagnostic features that may underlie these correlations, and suggested that dimensional classifications may better reflect the architecture of SMI than categorical labeling [3]. Despite these insights, many existing GWAS datasets are poorly suited for transdiagnostic investigations because they rely on structured diagnostic interviews built around branching logic. In such designs, symptoms are queried only within the context of specific diagnostic pathways, limiting the ability to systematically study symptoms that cut across diagnoses.

Here we report on analyses of the Paisa SMI Genetics Study, one of the largest SMI samples yet recruited from an admixed Latin American population. The Paisa population in northwestern Colombia descends from 16^th^-17^th^ century admixture of Indigenous American and Spanish founders and expanded from an initial bottleneck to a current size of ∼9 million [4–6]. This population is concentrated in mountainous regions and displays demographic and genetic characteristics typically associated with recently expanded genetic isolates [5–8].

The Paisa study includes 7,293 cases and 1,373 controls recruited in the Paisa region of Colombia, with data from diagnoses, transdiagnostic symptoms, and cognitive traits. We previously presented phenotype-only results in an initial subset of study participants [6]. The sample described here is underpowered for discovering SMI-associated genetic variants but well powered for analyses using Polygenic Risk Scores (PRS) of SMI GWAS to evaluate genetic relationships between phenotypes. Several studies have employed PRS for this purpose, for example, Ruderfer et al. showed that BD patients with psychotic symptoms carried higher PRS_SCZ_ than BD patients without psychotic symptoms, while SCZ patients with manic symptoms carried higher PRS_BD_ than SCZ patients without manic symptoms [9]. Similarly, Song et al. demonstrated that BD patients in remission had lower PRS_MDD_ and PRS_SCZ_ than BD patients not in remission, whereas BD patients with mood-episode psychosis had elevated PRS_SCZ_ compared to BD patients without mood-episode psychosis [10].

Such studies have been limited to European-ancestry populations, however, and have not systematically examined the effect of diagnosis-based PRS on cognitive performance. In the analyses reported here we applied both traditional and local ancestry-specific genetic association models to measure population-specific contributions to SMI-related phenotypes. By integrating diagnosis PRS, symptom-level traits, and cognitive domains, we (i) quantify the impact of ancestry on PRS accuracy and predictability in an admixed population, (ii) characterize transdiagnostic associations with symptoms and cognitive traits, and (iii) identify ancestry-specific associations in SMI loci. Our analyses highlight the overlap of loci shared across all SMI diagnoses, and associations with the cognitive domains of executive function and social cognition, and psychosis-related symptoms. They demonstrate the value of studying admixed populations while emphasizing the need for larger non-European cohorts.

## Methods

### Sample Ascertainment

The Paisa SMI Genetic Study recruited SMI cases (agnostic to specific diagnosis) and control participants between 2017 and 2022 in two departments (states) of the Paisa region. To enrich the sample for members of the Paisa population isolate [5], we recruited only individuals with both paternal and maternal surnames characteristic of this population [4, 11]. Cases were recruited from three hospitals: Clínica San Juan de Dios de Manizales (CSJDM), Hospital Universitario San Vicente Fundación (HUSVF), located, respectively, in the departments of Caldas and Antioquia and the Hospital Mental de Antioquia (HOMO), a public psychiatric hospital. Criteria for study inclusion were the same as reported previously [6]. Cases were age ≥18 with no known first-degree relative already enrolled and required a clinician-confirmed diagnosis of severe mood or psychotic disorders; exclusionary criteria included substance-induced presentations, neurological conditions, or intellectual disability. Controls were drawn from the same communities as cases and were screened to exclude severe psychiatric or substance use disorders. All participants provided informed consent, and study protocols were approved by institutional review boards in Colombia and the U.S. (UCLA IRB#16-002084). A detailed description of recruitment and sample ascertainment is in the Supplementary Material.

### Study Measures

As previously [6], lifetime diagnoses and cross-diagnosis symptom-level data were obtained through structured interviews using the Spanish NetSCID, a computerized version of the Structured Interview for DSM-5 [12, 13]. To assess cross-diagnostic symptomatology in cases, we administered seven supplementary questions about fatigue, grandiosity, decreased need for sleep, flight of ideas, hypersomnia, apathy, and anhedonia. We encoded symptoms as present if the participant endorsed them at any point in the interview, and used the information obtained in the supplementary questions to fill in data missing due to the branching nature of the NetSCID. A total of 14 symptoms were analyzed in this study, including the seven mentioned above and depressed mood, avolition, suicidal thoughts, suicide attempt, any delusion, any hallucination, disorganized speech, and grossly disorganized behavior. For descriptive purposes we classify these symptoms into one of three domains (depression, mania, and psychosis) based on results of an item-factor analysis that we reported previously [6]. These symptoms were selected as they are transdiagnostic [14], and may capture both shared genetic liability [15–17] and dimensional components of SMI [18].

We assessed speed and accuracy of neurocognitive performance across domains that we previously found to be associated with SMI [6] (executive function, memory, complex cognition, social cognition, and motor speed), using nine tests from the Penn Computerized Neurocognitive Battery (CNB) [19–21]. Data for speed were multiplied by –1 so that poorer performance (longer response time) would result in a lower value. Age, sex, years of education, and the interaction between education and age strongly affect neurocognitive performance; therefore, the raw data for each test were regressed on sex, age, age^2^, age^3^, and interaction of education and age [22]. Residuals from this regression were inverse-normal transformed and used for further analyses.

All study data were collected and managed using REDCap electronic data capture tools hosted at UCLA [23, 24]. Statistical analyses used R v4.1.0 and v4.3.1 [25].

### Genetic Data

Genotypes were obtained by DNA sequencing using the Blended Genome Exome (BGE) technology, in the Populations Underrepresented in Mental Illness Associations Study (PUMAS). BGE is a DNA library blending approach that generates low coverage (1-4x) whole genome sequences and deep whole exome sequences (30-40x) [26]. The sequenced samples were imputed using the Genotype Likelihoods Imputation and Phasing (GLIMPSE2) method with the HGDP+1kGP reference panel (paper in press) [27].

We removed samples that failed sequencing, with outlier values (defined as values >8 median absolute deviations from the mean for the following metrics: N singletons, N insertions, N deletions, N transitions, N transversions, heterozygosity ratio, transition to transversion ratio, and insertion to deletion ratio), or for which self-identified birth sex did not match genotype-defined sex.

### Principal Components Analysis

We constructed a genetic relationship matrix (GRM) using LD-pruned, common (frequency >1%) variants. Variants in the MHC region were removed prior to analysis. We used this GRM to estimate the top 20 principal components (PCs). Both construction of the GRM and estimation of the PCs used GCTA [28]. Prior to conducting the PCA, we used NAToRA [29] to prune the sample to a set of participants maximally related at the level of first cousin. After PCA was complete, we projected the excluded individuals back into the component space using PCA factor loadings [30]. Subsequent analyses included all participants, using the GRM to account for relatedness.

### Ancestry: Estimation of Global and Local Admixture Proportions

We used the ADMIXTURE program [31] and European (EUR, n=752), African (AFR, n=992), and South/Central American (AMR, n=191) HGDP reference samples to estimate global admixture proportions. Multiallelic SNPs and A/T or G/C SNPs were excluded. The analysis used LD pruned variants with MAF>1% that were present in these three reference sets and our Paisa sample.

To calculate local ancestry, we followed a published pipeline [32] with minor changes. Briefly, BGE genotype data were phased using EAGLE v2.4.1 [33] and local ancestry was estimated from the phased files using FLARE v0.5.2 [34]. For computational efficiency, FLARE’s pipeline filters out SNPs with redundant ancestry information and low frequency. We changed MAF setting to 0.01 to minimize the loss of low frequency variants, and we interpolated the local ancestry of excluded SNPs. The ancestral allele information was drawn from the same HGDP reference panel used in the global admixture analysis. The ancestry annotated files were further processed with Tractor v1.4.0 [35] to generate *hapcount* and *dosage* files which indicate for each SNP/ancestry both the number of ancestral alleles and the number of minor alleles carried on a given ancestral haplotype. To evaluate the consistency of the local ancestry inference, we estimated the global ancestry proportions, for each individual, as the count of each type of ancestral allele divided by the total allelic counts, and compared these estimates to global admixture proportions derived from ADMIXTURE.

### Analytic Strategy

To examine the impact of known GWAS signals in the Paisa sample, we analyzed both PRS and SNP-level associations. We evaluated: i) the ability of the PRS to predict SMI diagnoses, symptoms, and cognitive traits; ii) the effect of ancestral background on the variability of individual PRS, iii) SNP-level associations to phenotypes in loci with significant associations to SMI in published GWAS from other study populations.

### Polygenic Score Estimates

We generated PRS for four SMI diagnoses [SCZ, BD, BD type I (BD1), MDD], using summary statistics from the most recent published GWAS from international consortia for which data are publicly available [2, 36–38]. Mirroring the GWAS on which the PRS are based, we separated BD1 from BD overall, as the latter designation includes individuals with both BD1 and a milder form of BD (BD2); while BD1 is more strongly genetically correlated with SCZ than with MDD, BD2 is more strongly genetically correlated with MDD than with SCZ [39]. We also generated PRS for reaction time (RCT), as it is the only measure of general cognitive function with available GWAS summary statistics from a large cohort (UK Biobank) and is phenotypically similar to the Digit Symbol Substitution Test with the matching paradigm (sometimes referred to as Digit Symbol Match [6]) assessed in the Paisa sample [40]. We included PRS for height as a negative control; it is a phenotype for which GWAS data are available on large diverse samples and demonstrates minimal genetic correlation with SMI phenotypes [41].

The GWAS summary statistics were remapped from hg19/GRCh37 to GRCh38 when necessary, harmonized to our dataset, and restricted to biallelic SNPs. Polygenic scores were calculated with PRScs [42] (auto mode, 10,000 iterations, 500 burn-in) using the UK Biobank European LD reference panel, and scores were computed with PLINK [43]. To create ancestry-specific partial PRS, ancestry-specific dosages were aligned to the same alleles used in the PRScs weights. These dosages were then multiplied by the corresponding allele-specific GWAS effect sizes and summed genome-wide to compute ancestry-specific polygenic scores for each diagnosis, following a standard PRS framework. This procedure resulted in one polygenic score for each ancestry, which we denote ancestry-specific partial polygenic scores (pPRS_AFR_, pPRS_EUR_, pPRS_AMR_).

### Polygenic Score Analyses

To evaluate PRS variability we used the individual accuracy principle [44], which uses individual variability in PRS to estimate the accuracy of PRS estimates; people who are genetically distant from the reference population show higher polygenic score variance, and in turn lower PRS accuracy, than those genetically close to the reference population. Using this approach, we derived individual polygenic score variance from the posterior samples produced by PRScs and use ancestry as a proxy for genetic distance to the predominantly European training data.

The ability of the PRS to predict diagnoses, symptoms, and cognitive traits was evaluated using regression models. We first regressed the PRS on the first four PCs, and estimated the residuals (R) from this regression:

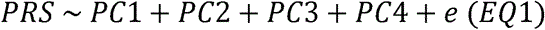

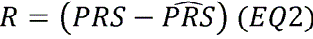

where *e* is residual error. We used four PCs as we found only the first four PCs to be related to global ancestry, and inclusion of additional unrelated PCs could run the risk of inflating the false positive rate [45]. The residuals were standardized (SR) to Z-scores based on the mean and standard deviation (SD) of the control group (Ctrl).

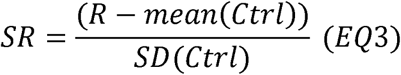

The SR were used as predictors in logistic regression analyses to predict the binary outcomes of diagnosis and symptom presence/absence. A separate sensitivity analysis was done where we did not regress out the PCs from the PRS but included them in the model of EQ4 as a fixed effect.

To account for the high degree of cryptic relatedness in the Paisa population, we used mixed-effect logistic regression models, incorporating the GRM to account for related individuals.

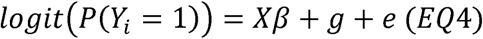

where *Xβ* are fixed effects, 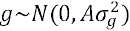 is the polygenic random effect with A being the GRM and *e* is residual error. In the analysis of diagnosis as an outcome, Y_i_ is 1 for cases, and 0 for controls, and *Xβ* contains the intercept term and SR. In the analysis of a symptom as an outcome, Y_i_ is 1 for cases (of any diagnosis) with the symptom, and 0 for cases (of any diagnosis) without the symptom (controls were not assessed for presence/absence of symptom data), and *Xβ* contains the intercept, SR, and the categorical diagnosis. We include diagnosis as a covariate in the logistic regression of symptoms in cases, because while most of the symptoms are transdiagnostic (occurring in multiple different diagnoses), symptoms are more prevalent in some diagnoses than others and we wanted to estimate the impact of the PRS on symptom after accounting for the effect of diagnosis on symptom.

To analyze the quantitative cognitive traits as outcomes, EQ4 becomes a linear regression, and uses both cases and controls with quantitative data. The outcome is an inverse normal transformed residual quantitative trait, prepared as discussed above under “Study Measures” and *Xβ* contains the intercept, SR, and the categorical diagnosis. We include diagnosis as a covariate in the linear regression because there are known differences in cognition between different diagnoses[6], and we wanted to estimate the impact of the PRS on cognitive traits after accounting for the effect of diagnosis on these traits. For all analyses, since the PRS were standardized to Z-scores with controls as reference, the regression coefficients represent the effect of a one SD increase in PRS relative to controls.

Lastly, we employed simulations to evaluate if the loss of predictability of ancestry-specific partial PRS, compared to the full PRS, could be due to the size of AFR and AMR partial PRS (number of SNPs of that specific ancestry in the genome), or due to the content of the PRS in terms of non-EUR ancestral background. For each non-EUR ancestry (AFR and AMR), we generated simulated partial PRS (pPRS_AFR_*, pPRS_AMR_*) by randomly permuting the SNP positions originally annotated to that ancestry within each individual. At these permuted locations, we used the EUR genotype dosages and their corresponding effect sizes to construct a simulated partial PRS. This step produced simulated scores using the same number of SNPs as the observed pPRS. We generated 100 replicates and used them as a null benchmark to the observed pPRS. We expected that if the loss of prediction was due simply to the number of SNPs used in the construction of the pPRS, the simulations would perform similarly to the real observed pPRS. In contrast, if the loss of prediction is due to the content, the simulated PRS will perform better than the observed PRS, as the decreased performance due to unmatched ancestry background would be partially corrected.

### Association to Paisa phenotypes at SMI loci from published GWAS

To evaluate associations, in the Paisa sample, of SNPs shown in prior studies to be associated at genome-wide significance to SMI diagnoses, we formed these SNPs into loci using the published GWAS summary statistics [2, 36–38]; for each GWAS SNP we identified the minimum GWAS p-value across all external GWAS in which it was analyzed, and then used the resultant set of p-values in PLINK’s “clump” procedure (with index SNP p-value threshold = p<5×10^−8^, secondary significance threshold = p<1×10^−5^, r^2^=0.10 to the lead SNP [using LD data from the Paisa sample], and physical distance=250kb) [43]. Each resulting locus was labeled with a diagnosis (BD, BD1, MDD, or SCZ) if at least one of the variants in the locus was associated at p<5×10^−8^ in the respective GWAS. This procedure resulted in some loci being labeled for more than one diagnosis. We included SNPs with 1×10^−5^ < p < 5×10^−8^ that were in LD with the lead SNP at each locus in order to account for possibly different LD patterns in Paisa compared to the GWAS samples.

Marginal associations between each SNP in these loci to each phenotype (diagnoses, symptoms, and traits) in the Paisa sample were explored using mixed effects linear (quantitative traits) or logistic (diagnoses and symptoms) regressions as implemented in SAIGE. The first four PCs were included as covariates, and the GRM described above was used to account for relationships among participants [46]. Ancestry-specific associations were estimated using Tractor v1.4.0 [35], which models the effect of risk alleles conditional on their ancestral background by adjusting for local ancestry at each haplotype, including the first four principal components as covariates. Significant associations identified in the Tractor analysis were subsequently re-evaluated using a mixed model, incorporating a genetic relationship matrix (GRM) to account for cryptic relatedness.

For analysis of diagnoses, cases were participants with the indicated diagnosis, and controls were participants without any psychiatric diagnosis. Individual symptoms were not queried in control participants; therefore, when analyzing symptom presence/absence we employed the mixed model in cases with and without the symptom, irrespective of diagnosis, and did not include control participants. As symptom prevalence and cognitive trait means differed among diagnostic categories, we conditioned these models by diagnoses, in addition to the first four PCs.

To adjust the significance threshold for multiple testing, we used a Bonferroni correction based on the number of loci tested for standard associations, and three times this number for ancestry specific associations (adjusting for testing AFR, AMR, and EUR ancestries); we did not adjust for multiplicity across traits. These thresholds serve as an instrument to summarize our findings, rather than a guarantee of significance. In addition, to evaluate the overall signal strength, we used the Kolmogorov Smirnov and Higher Criticism statistics to measure the distance between the distribution of p-values for association tests (across loci and across phenotypes) and the uniform distribution, which is expected when all of the tested hypotheses are null.

### Replication of diagnosis-associated loci

To assess the replication in our dataset of published genome-wide significant SNP associations to the SMI diagnoses, we examined concordance of effect direction. We used an exact binomial test (one-sided) in independent variants (the most significant variant in each locus, as estimated from the published GWAS) to evaluate the null hypothesis of randomly oriented direction of effects. Rejection of this null would indicate that the direction of effect in Paisa and the original GWAS was the same more often than expected by chance.

## Results

### Characterization of the genotyped study sample

We recruited 9,107 participants; following quality control of the sequence data we analyzed a genotyped study sample consisting of 8,666 participants (1,373 controls and 7,293 cases, recruited using a uniform approach to select case participants agnostic to SMI diagnosis (Methods, see Table 1, Supplementary Table 1). The proportion of cases with diagnoses of SCZ, BD, and MDD differ between the clinical sites (Supplementary Figure 1), likely reflecting the different patient populations served by HOMO (where the most frequent diagnoses are SCZ and BD1) and CSJDM (where the most frequent SMI diagnosis is MDD).

**Table 1.** Clinical and ancestry variations in the genotyped study sample, by recruitment site.

|  | Site |  |
| --- | --- | --- |
|  | Caldas<br>n= 5,283 | Antioquia<br>n= 3,383 |
| Age: mean (min-max) | 42.21 (18-86) | 46.00 (18-85) |
| Sex: Female (%) | 3,345 (63.32) | 1,831 (54.1) |
| **Diagnosis** |  |  |
| Control | 791 (15.00) | 582 (17.20) |
| MDD | 2,335 (44.20) | 327 (9.67) |
| SCZ | 297 (5.62) | 829 (24.50) |
| BD1 | 1,004 (19.00) | 1,294 (38.32) |
| BD2 | 450 (8.52) | 100 (2.96) |
| SCZA | 94 (1.78) | 209 (6.18) |
| Other BD | 288 (5.45) | 22 (0.65) |
| Other SMI diagnosis | 24 (0.45) | 20 (0.59) |
| BD (all BD) | 1,742 (39.45) | 1,416 (41.93) |
| Other (SCZA+ Other psychosis) | 118 (2.23) | 229 (6.77) |
| **Ancestry** |  |  |
| AFR: mean (SD) | 0.05 (0.05) | 0.11 (0.08) |
| EUR: mean (SD) | 0.59 (0.10) | 0.57 (0.12) |
| AMR: mean (SD) | 0.36 (0.08) | 0.32 (0.08) |

Cognitive performance scores were recorded for 5,566 participants with BGE data (4,473 cases and 1,193 controls, Supplementary Table 2). As in our earlier work [6], MDD patients and controls showed similar scores for both accuracy and speed in all tests; these scores were higher than those observed for BD1 and SCZ patients (which were similar to each other) (Supplementary Figure 2).

Global estimates (Table 1) indicate that EUR ancestry is the largest contributor to this sample (mean = 58.02%, SD = 10.46) followed by AMR (mean = 35.54%, SD=7.82), and then AFR (mean=7.44%, SD=7.02). The Caldas and Antioquia samples differed most strikingly in the frequency of AFR ancestry (0.05 vs 0.11, respectively). These proportions are comparable to those reported previously for Paisa region samples[4, 47, 48]. Principal components (PCs) up to the PC4 capture most of this ancestry-associated genetic variability, with PC1 and PC2 effectively separating ancestry in a continuum across 3 ancestries (Figure 1).

**Figure 1.**
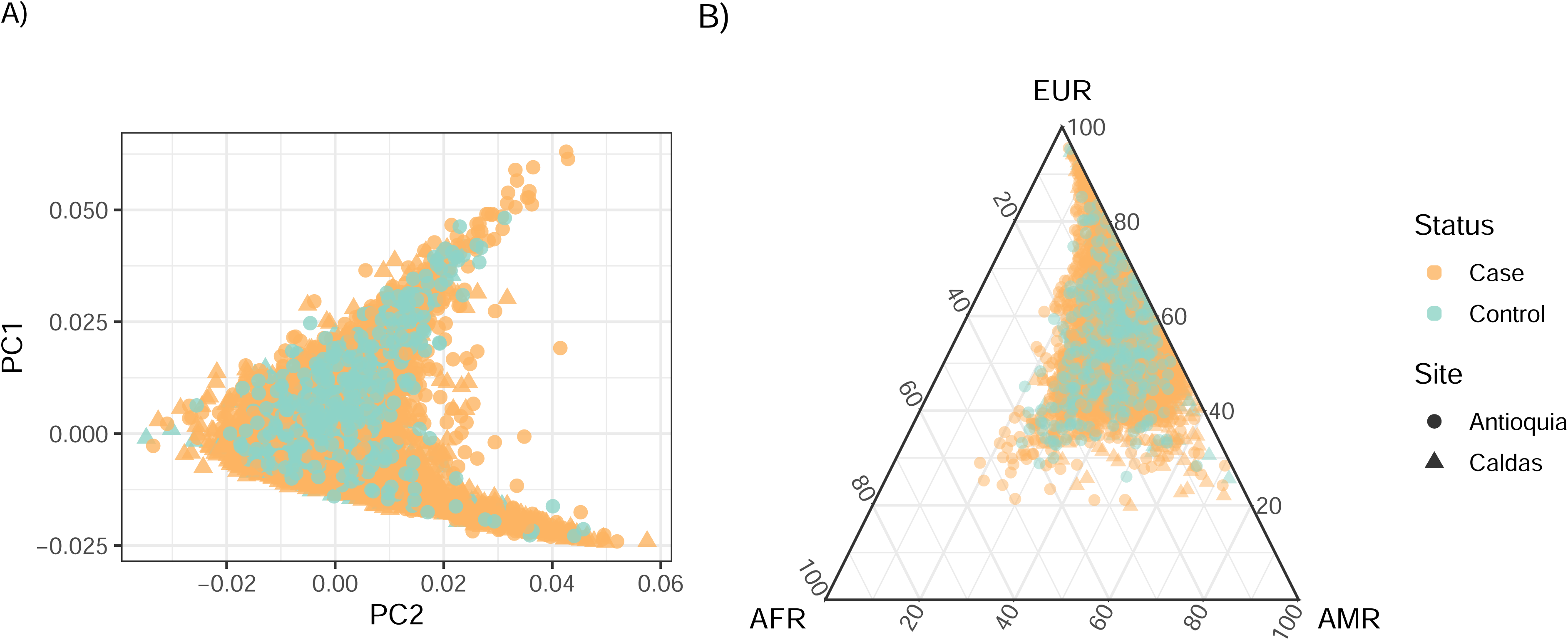
Distribution of the first two principal components (A) and global ancestry proportions (B) in the Paisa sample. Points represent an individual, color-coded by case/control status. Antioquia: Hospital Universitario San Vicente Fundación (n = 303) and HOMO: Hospital Mental de Antioquia (n= 3,080). Caldas: Clínica San Juan de Dios de Manizales (n= 5,283).

### Polygenic scores from psychiatric GWAS are predictive of diagnoses in the Paisa

For each set of GWAS summary statistics (Supplementary Table 3), approximately 7.5 million SNPs were remapped to GRCh38, and 6,631,771 SNPs across all the GWAS were harmonized to the SNPs in the Paisa BGE data (Supplementary Table 4). All four diagnosis-based PRS predicted their respective diagnoses in the Paisa sample (Figure 2, Supplementary Table 5). The PRS for schizophrenia (PRS_SCZ_) showed the strongest degree of prediction; the odds of SCZ for SCZ cases increased 2.09 for each one-unit SD unit increase in PRS_SCZ_ (95% CI: 1.89–2.30, p = 7.72×10^−51^) relative to controls, a result consistent with the fact that SCZ has a higher SNP heritability than the other diagnoses and that the GWAS on which its PRS is based was better powered than the GWAS of these diagnoses [49, 50]. The PRS_SCZ_ also predicts BD and BD1 as effectively as their respective PRS, while both the PRS_BD_ and the PRS_BD1_ each performed better in predicting BD1 than in predicting BD overall. The PRS_BD,_ PRS_BD1_ and PRS_SCZ_ performed almost as well in predicting MDD as did PRS_MDD_ itself. Neither the PRS_RCT_ nor the PRS_height_ were predictive of any of the psychiatric diagnoses, nor of the affected status comprising all diagnoses together. The sensitivity analysis including the PCs in a joint analysis with the PRS, rather than regressing out the PCs from the PRS prior to analysis (see Methods) produced nearly identical results (Supplementary Table 5).

**Figure 2.**
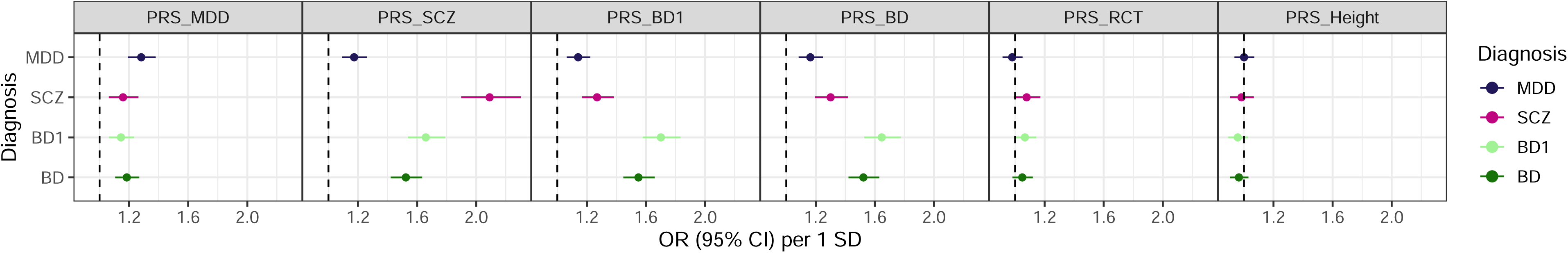
Association of polygenic scores across the diagnoses. Forest plot depicting odd ratios (OR) of the logistic regression of each diagnosis on the polygenic scores (MDD, SCZ, BD1, BD, Height, RCT). Polygenic scores were calculated from external GWAS. The points represent the OR for each 1 standard deviation (SD) increase from the mean PRS of controls, and the error bars represent 95% confidence intervals. The vertical dashed line represents OR=1. PRS: polygenic risk score. BD: bipolar disorder (dark green), BD1: bipolar disorder type I (light green), SCZ: schizophrenia (magenta), and MDD: major depression disorder (blue).

The four diagnostic PRS are correlated with each other (Supplementary Table 6). As a sensitivity analysis we performed multivariate prediction of each diagnosis, using PRS_SCZ_, PRS_MDD_, and PRS_BD1_ (we omitted PRS_BD_ because of its strong correlation with PRS_BD1_). For SCZ the predictive power of multiple PRS seems to be due to common signal, e.g., the predictive contributions of PRS_MDD_ and PRS_BD1_ diminish when PRS_SCZ_ is included in the same model. In contrast, for BD1, the individual PRS appears to contribute more independent predictive signal (Supplementary Figure 3).

### PRS precision and predictability are maintained at reduced levels in a non-reference ancestral background

To evaluate the accuracy of diagnosis-based PRS in the Paisa population, we examined individual-level PRS variability across ancestry proportions and estimated global predictive performance using ancestry-specific partial PRS compared with simulated scores. Individual PRS variance increased with increasing AFR ancestry proportions (r=0.22, p=12.99×10^−130^) and decreased with the proportion of EUR ancestry (r=-0.19, p=9.79×10^−7^, Supplementary Figure 4); this result is expected when using an EUR reference to estimate the PRS in an admixed population [44].

The predictive performance of partial PRS across all diagnoses followed a consistent gradient, with the highest prediction presented by the pPRS_EUR_ estimates, slightly reduced prediction for pPRS_AMR_ estimates and the lowest prediction for pPRS_AFR_ estimates, reflecting not only mean global ancestry proportions in our sample, but also the decrease of power with the increase of genetic distance from the European reference population (Supplementary Figure 5). This gradient in prediction ability is more pronounced in SCZ, where pPRS_EUR_ predicts 50% more risk of SCZ than the pPRS_AFR_, and in BD1 where pPRS_EUR_ predicts 46% more risk of BD1 than pPRS_AFR_. For MDD, the gradient was detectable, but effect sizes were smaller overall, while in BD the pattern resembled BD1 with slightly attenuated effects (Supplementary Table 7).

Simulation analysis showed that for BD, BD1 and SCZ the simulated AFR-specific pPRS consistently outperformed the real observed pPRS_AFR_ in predictive models (Figure 2). The average odds ratio across simulations was 1.12 (95% CI: 1.06–1.18) for BD1, 1.11 (95% CI: 1.06–1.16), and 1.17 (95% CI: 1.10–1.24) for SCZ, and for all of them >88% the simulated odds ratios exceeded that of the real observed score. This result indicates that the low prediction of the observed pPRS_AFR_ is not fully explained by number of SNPs used to calculate it; instead it is partially affected by lack of EUR background, as better prediction ability is achieved with random ancestral background than with the specific AFR background. In contrast, in MDD the point estimates of the pPRS_AFR_ simulations fall within the confidence interval of the real observed pPRS_AFR_ 96% of the times; therefore no improvement in prediction is observed. This observation could suggest that the MDD GWAS is not as homogenous as was reported. The behavior of the AMR-specific partial polygenic scores was not the same; simulation did not show an improvement in prediction of BD1 and MDD and had a poorer prediction for SCZ and BD1. This result is not unexpected, as the AMR ancestry is closer to EUR ancestry, and likely confounded by admixed AMR reference samples; this is a limitation difficult to overcome with currently available AMR data. (Supplementary Figure 6).

### Signals from SMI genetic associations are observed for transdiagnostic symptoms and cognitive traits

Most symptoms were observed at prevalences >10% across all diagnoses (Supplementary Table 8, Supplementary Figure 7). We observed three significant associations between PRS_SCZ_ and transdiagnostic symptoms (Figure 3, Supplementary Table 9). Specifically, a one SD increase of PRS_SCZ_ from the control mean was significantly associated with an increased risk of delusions (OR=1.12, 95%CI: 1.06-1.18, p=2.58×10^−5^) in cases, as has been observed previously in predominately European populations [51–53] and with a decrease in risk of both depressed mood (OR=0.89, 95% CI:0.85-0.95, p=1.93×10^−4^), and lifetime suicidal ideation (OR=0.89, 95% CI:0.84-0.94, p = 3.77×10^−5^, Figure 3, Supplementary Table 9), a result that was also observed previously in predominately European populations [54].

**Figure 3.**
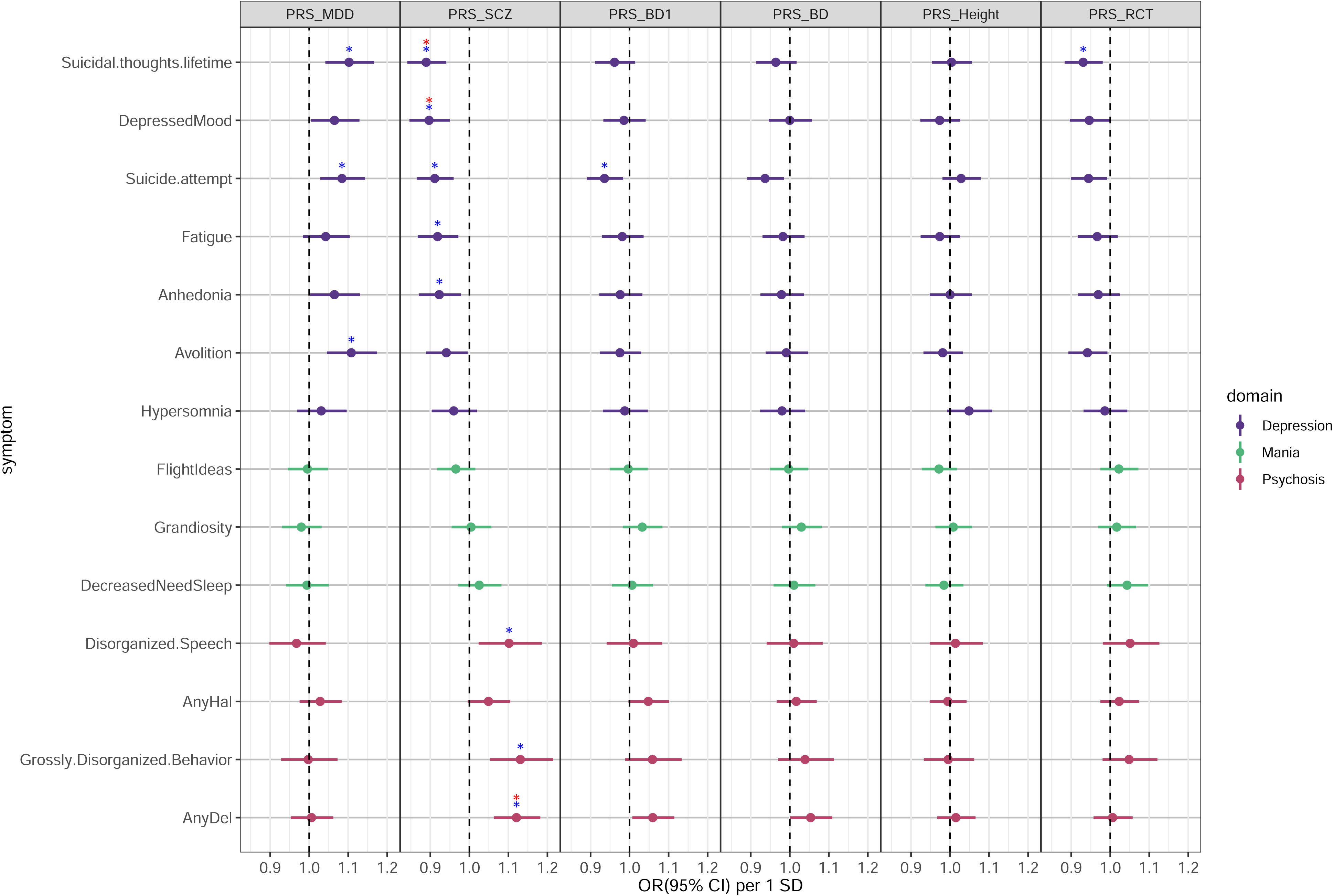
Association of polygenic scores of psychiatric diagnoses with psychiatric symptoms in cases. Forest plots indicating odds ratio (OR, points) and 95% confidence intervals (error bars) of the logistic regression model of diagnosis PRS on 14 transdiagnostic symptoms. PRS were calculated from external GWAS, and the first four PCs, and diagnosis was regressed out before application of logistic regression. Dashed lines indicate OR= 1.0. Associations under the multiple testing correction threshold (p < 3.23×10^−4^) are indicated by the red asterisk, associations under the FDR<0.05 threshold with the blue asterisk. AnyHal: Any Hallucination, AnyDel: Any Delusion.

PRS_SCZ_ was also significantly associated with cognitive measures in five distinct cognitive domains (Figure 4, Supplementary Table 10). A one SD increase in PRS_SCZ_ was associated with slower social cognition speed in the emotion identification test (EmoID, effect = −0.07, 95% CI: –0.09 to –0.04, p = 3.2×10□□) and the emotion intensity differentiation test (EmoDiff, effect = −0.06, 95% CI: −0.08 to −0.03, p = 6.65×10□□), findings consistent with prior results from predominately European populations [55]. Increasing PRS_SCZ_ was also associated with lower Digit Symbol Substitution Test accuracy and speed, matrix analysis speed, and digit recall accuracy. This finding aligns with previously reported negative correlations for cognitive traits for PRS_SCZ_ [56, 57]. Increasing PRS_BD1_ was associated with decreased speed of emotion identification (EmoID, effect = –0.051, 95% CI: –0.077 to –0.024, p=2.27×10□^4^). No cognitive trait was associated with the negative control PRS_height_. This negative control PRS was only associated with the negative control phenotype (weight), which in turn was not associated with any of the diagnostic PRS.

**Figure 4.**
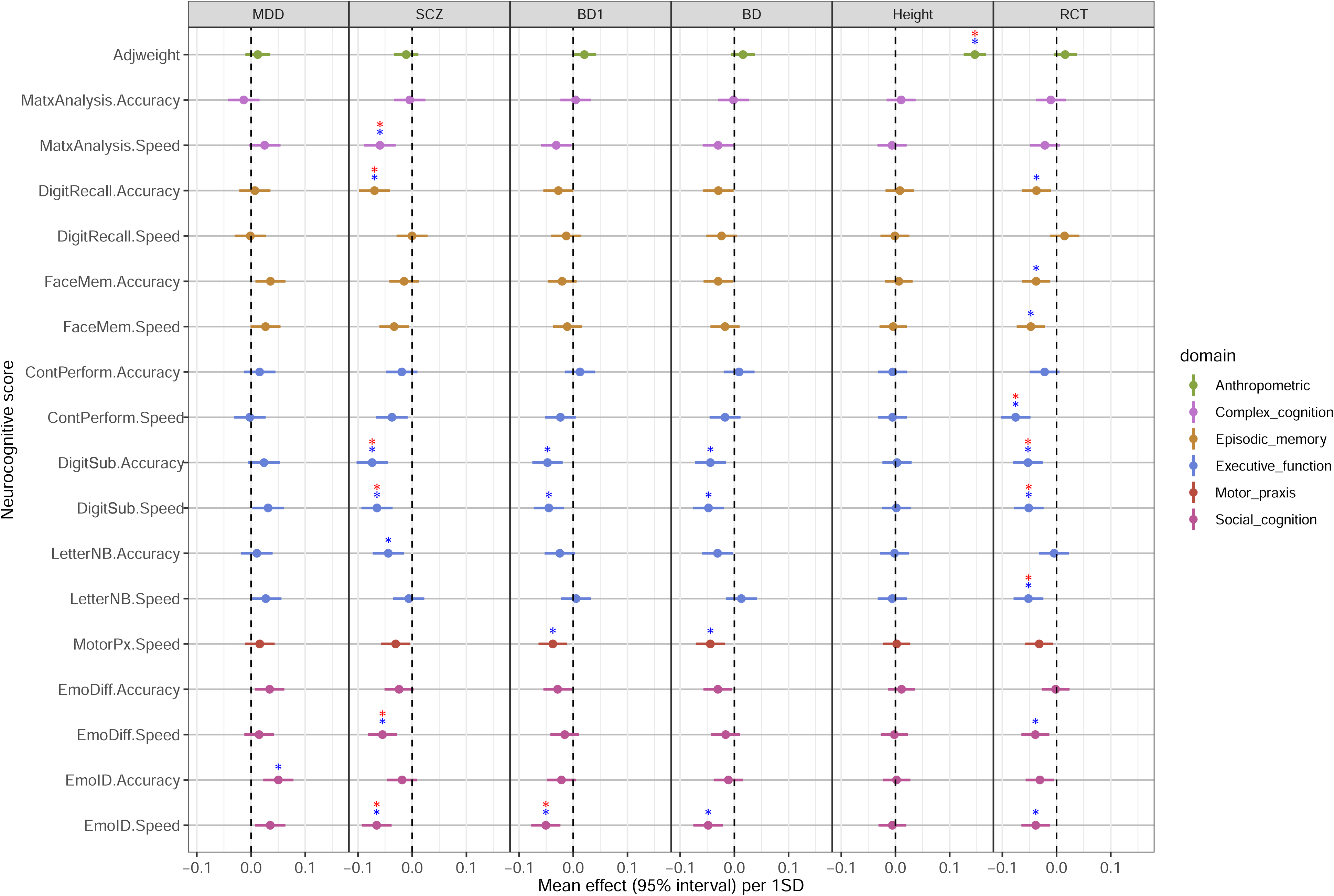
Association of polygenic scores of psychiatric diagnoses with cognitive performance scores in Paisa sample. Forest plots indicating effect size (points) and 95% confidence intervals (error bars) of the linear regression model of diagnosis PRS on 17 speed and accuracy scores of the Penn Computerized Neurocognitive Battery (CNB). PRS were calculated from external GWAS, and the first four PCs, and diagnosis was regressed out before application of linear regression. Dashed vertical lines indicate OR= 1.0. Associations under the multiple testing correction threshold (p < 3.23×10^−4^) are indicated by the red asterisk, associations under the FDR<0.05 threshold with the blue asterisk. MatxAnalysis: Matrix analysis test. DigitRecall: Digit symbol test, recall trials. FaceMem: Face memory test. DigitSub: Digit symbol substitution test. LetterNB: Letter-N-back test. ContPerform: Continuous performance test. MotorPx: Motor praxis test. EmoID: Emotional identification test. EmoDiff: Measured emotion differentiation test. SD: Standard deviation.

The PRS_RCT_ was not associated with any of the transdiagnostic symptoms but was significantly associated with four measures of cognitive executive function. A one SD increase in PRS_RCT_ (corresponding to slower speed and reduced cognitive performance) was associated with decreased overall accuracy (effect = –0.05, SE=0.01, p = 1.54×10^−4^) and speed (effect = –0.05, SE= 0.01, p =2.26×10^−4^), as well as with decreased speed in the letter N-back test (LetterNB, effect = –0.05, SE= 0.01, p =2.16×10^−4^), and the continuous performance test (ContPerform, effect = –0.08, SE = 0.01, p = 6.89×10^−8^, Figure 4, Supplementary Table 10). Three out of four associations are indicative of decreased speed (increased response time, and poorer cognition) with increasing PRS_RCT_, supporting the interpretation that reaction time PRS captures a genetic component of cognitive performance in individuals with SMI.

Lastly, there was no significant association of the ancestry pPRS with symptoms (Supplementary Figure 8a), but in the cognitive traits most of the associations that were significant with the traditional PRS were also observed in the pPRS_EUR_ at an FDR<0.05 (Supplementary Figure 8b). This result is consistent with the fact that discovery GWAS were primarily conducted in EUR samples.

### Identification of SMI-associated loci in the Paisa sample

We identified 692 independent loci that included 56,302 SNPs with p<1×10^−5^ from the four published SMI GWAS. Each locus included SNPs with p<5×10^−8^ in the original GWAS; SNPs with 1×10^−5^ < p < 5×10^−8^ were in LD with the GWAS lead SNP in the same locus. Of these 692 loci, 567 are labeled by the GWAS for one diagnosis, and 125 loci are labelled by GWAS for at least two diagnoses: 90 loci labelled by GWAS of two diagnoses, 26 loci labelled by GWAS of three diagnoses, and nine loci labelled by GWAS of all four diagnoses (Supplementary Table 11). In total, these loci include 274 MDD loci, 449 SCZ loci, 58 BD1 loci and 80 BD loci.

For all diagnoses, we observed more concordance in direction of effect than expected by chance: MDD (observed: 156/274, p_binom_=1.26×10^−2^), SCZ (observed: 317/449, p_binom_= 6.18×10^−19^), BD1 (observed: 41/58, p_binom_=1.1×10^−3^), and BD (observed: 54/80, p_binom_=5.20×10^−4^) (Supplementary Table 12). These results support a role for these loci in contributing to SMI risk in the Paisa sample.

### Diagnoses, symptoms, and traits show ancestry-specific distinct genetic associations at the locus level

We evaluated genetic associations with diagnoses, symptoms and cognitive traits in two models for the 56,278 SNPs with ancestry information (40,670 annotated, and 15,608 interpolated with the annotated SNPs; 24 SNPs could not be reliably interpolated) located in the 692 loci. We found a total of 433 SNPs in 48 loci that passed our significance threshold; 376 SNPs (in 28 loci) were significant in the traditional association model, and 155 SNPS (in 29 loci) were significant in the ancestry specific Tractor model; 55 SNPs (in nine loci) had associations in both models (Supplementary Tables 13, 14, and Figure 5).

**Figure 5.**
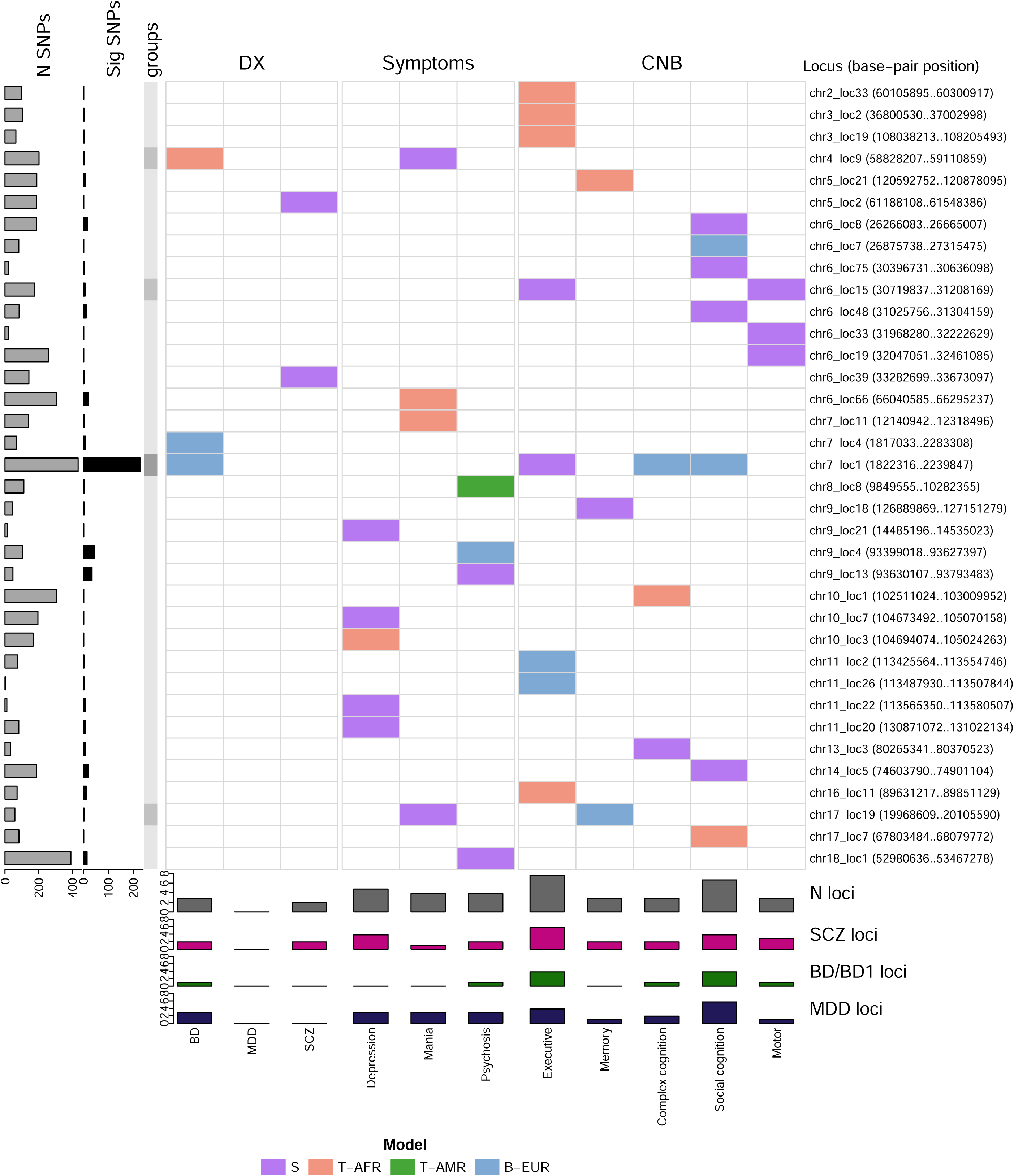
Overview of significant genetic associations by locus. Summary of the loci with more than one SNP associated (with a p-value < 7.23×10^−5^ in the SAIGE model or p-value < 2.41×10^−5^) with phenotypes in the domains of diagnosis, symptoms or cognitive traits (CNB). Colored cells indicate an association is present for the corresponding locus (right column). Bars in the bottom indicate the number of loci associated for the corresponding domain, the top bars (grey) indicate total number of loci, and the colored bars correspond to the number of loci labelled as significant in the corresponding published GWAS (loci labelled for BD and BD1are counted together). N SNPs: number of SNPs analyzed in the locus. Sig. SNPs: Number of analyzed SNPs below the corresponding significance threshold. S: SAIGE significance, T-AFR: African specific Tractor significance, T-AMR: Admixed American specific Tractor significance, B-EUR: both SAIGE and European specific Tractor significance.

The 48 significant loci involved 30 of the 35 investigated phenotypes, for a total of 57 locus-phenotype associations, 31 of which corresponded to ancestry specific associations (18 to AFR, 12 to EUR, 1 to AMR). We did not observe any locus with ancestry specific associations to more than one ancestry. The high abundance of AFR-specific associations observed in the Paisa SMI sample here likely reflects several features that characterize populations with substantial levels of AFR ancestry: higher genetic diversity, shorter LD blocks, and greater allele frequency differentiation relative to EUR. All these features increase Tractor’s ability to detect ancestry-specific associations [35]. In contrast, AMR ancestry is genetically closer to EUR, more confounded by admixture, and less variable, which reduces Tractor’s ability to identify AMR-specific effects. EUR effects are already well captured by the discovery GWAS, so Tractor provided less additional power for detecting EUR-specific associations.

Most significant loci are associated with only a single trait, while five loci showed associations with more than one phenotype in either model (Figure 5). Notably, one locus, on chromosome 7 showed associations with four phenotypes in our sample (BD, executive function, complex cognition, and social cognition). The analysis of the global signal (Supplementary Figure 9) confirmed that these associated loci carry substantial information for the traits under study, reproducing, at a more granular level, the results obtained with PRS. This observation underscores the considerable portability from EUR to AMR samples, and shared genetic signatures across traits. Additionally, it is notable that the overall association signal between these 692 loci and the cognitive traits is comparable to the signal from the diagnoses which were originally used to identify the loci.

## Discussion

This study expands our previous efforts to genetically characterize SMI in an admixed Latin American population sample with detailed clinical and cognitive phenotyping. The dataset analyzed here is more than triple the size of that in our prior phenotype-only analyses and now includes over 3,000 case participants from the largest public psychiatric hospital in Colombia. Analyses of genome-wide genotypes in the dataset show that genetic association signals identified in EUR populations for SMI diagnoses and for a cognitive trait (reaction time) are also observed in the admixed Paisa population; the portability of these signals is more evident at the level of PRS than at the locus level. These signals are observed in our dataset for SMI diagnoses, transdiagnostic symptoms, and cognitive phenotypes.

Our observation that PRS discovered in EUR ancestry samples are portable to the Paisa likely reflects the fact that, as in many admixed Latin American populations, the European contribution to Paisa ancestry is over 50% of the total. This result is consistent with the expectation that PRS performance decreases when calculated in populations that are farther from the one in which the associations were detected. There is a clear gradient in loss of predictability within the Paisa sample, from EUR to AMR to AFR, and future studies in this population will be facilitated by the development of improved references for both non-European ancestries.

We previously demonstrated that most individual symptoms are observed to differing degrees across all three main SMI diagnoses and that cognitive phenotypes stratify the overall Paisa SMI population. The work reported here supports those observations, showing that PRS from disease-specific GWAS were associated with symptom dimensions and cognitive traits. The disorder-specific PRS scores related well to the respective diagnostic categories, especially the PRS_SCZ_, consistent with the higher SNP heritability of schizophrenia compared to the other disorders. The correlations with clinical findings underscore the trans-diagnostic effects of genetic variation. Indeed, PRS_SCZ_ was as good a predictor of BD as the PRS targeted for BD. Regarding neurocognitive domains, social cognition, memory, aspects of complex cognition and processing speed stand out as phenotypes most strongly associated with PRS_SCZ._ These domains reflect the functioning of temporolimbic and fronto-parietal systems implicated in schizophrenia [58], and deficits in them are associated with severity and outcome in psychosis [59–61]. Our findings also align with recent recommendations for systematic measurement of social cognition in BD [62].

Taken together, the consistent associations between diagnosis-derived PRS and both symptom dimensions and cognitive performance suggest that a shared genetic signature is not only present across diagnoses, but maps onto specific clinical and cognitive features. While prior studies have demonstrated genetic overlap across psychiatric disorders [1, 3, 15–17], our findings extend this work by showing how this shared genetic signature relates to these domains within the same sample, and in an admixed Latin American population.

A similar pattern is seen at the locus level, where we identified transdiagnostic signal within the Paisa for multiple symptom domains and cognitive traits at five loci highlighted in GWAS as being associated with specific diagnoses. The strongest and broadest such signal was at chromosome 7p22.3, a locus that conferred increased risk of BD and was associated with reduced complex cognition and social cognition previously labelled as significant in GWAS of SCZ, MDD, BD, anorexia nervosa, attention deficit and hyperactivity disorder (ADHD) and autism spectrum disorders [63]. Additionally, this location gave one of the most significant findings, genome wide, in a multivariate GWAS for a combined SCZ-BD genomic factor [3]. Taken together, these findings suggest that 7p22.3 variation plays a role in cognition and in psychiatric disorders, transdiagnostically.

### Limitations and next steps

The Paisa sample is underpowered for genome-wide discovery of variants associated with SMI diagnoses or dimensional phenotypes. The cross-sectional design limits our ability to examine temporal relationships between genetic risk, symptom expression and cognitive performance. Because we did not assess symptoms in controls, we cannot determine if differences (in PRS or allele frequency) between cases with and without a symptom reflect overall SMI risk or a factor specific to that symptom. Finally, our findings should be interpreted with caution as they have not yet been replicated in independent cohorts.

Our results suggest the importance of future work focused on replications in independent cohorts, ancestrally diverse populations [64], and longitudinal studies; the latter effort would characterize how genotypes relate to changes in symptoms and cognition over time, and could be undertaken through genetic analyses of SMI-related phenotypes derived from EHR-linked biobanks in the Paisa region.

## Consent for publication

All participants who joined the studies listed under the above IRBs consented to the de-identified use of their data for general research use. During the consenting process, study staff explain the research goals, risks and benefits. Participants are also given the option to opt out of studies if they choose. Samples used in this research are all de-identified and have all been fully consented for inclusion in the study and any resulting publications.

## Availability of data and materials

All data generated and analyzed during this effort were used for Research and Development only. These data are not publicly available yet but will be available through NIMH Data Archive (NDA).

## Acknowledgements

The authors thank the CSJDM, HUSVF, HOMO, and all study participants for making this work possible.

## Author Contributions

MCR, JCM, JV recruited and interviewed participants and administered test instruments. TT, MMU, AMR, AMDZ, JValdez designed data bases, coordinated data transfers and managed data downloads. ELM, SKS and CS were responsible for data analysis. RCG, TMM oversaw production and analysis of cognitive data. SC and BMN oversaw production of sequence data. LOL and JFDH provided EHR data. JIE, VIR, CLJ, RCG, CEB, LOL, and NBF designed the study. ELM, SKS, CS, and NBF wrote the paper.

