## SupplementaryText for "Effect of ancestry and shared genetic architecture of serious mental illness on symptoms and cognition in an admixed Latin American population"

**Supplementary Figure Legends:**

**Supplementary Figure 1. Prevalence of diagnosis in the different recruitment sites.** Bars represent the proportion of every diagnosis relative to in each recruitment site. Antioquia: Hospital Universitario San Vicente Fundación (n = 303) and Hospital Mental de Antioquia (n= 3,080). Caldas: Clínica San Juan de Dios de Manizales (n= 5,283). HOMO: hospital mental de Antioquia. BD: bipolar disorder (includes BD1, BD2, and other BD). MDD: Major depression disorder, SCZ: Schizophrenia, Other: includes schizoaffective disorder type A, delusional disorder and brief psychotic disorder.

**Supplementary Figure 2. Prevalence of symptoms in the different diagnoses.** Bars represent the proportion of individuals with the diagnosis presenting each symptom. Colors indicate the factor domain most correlated with the diagnosis according to previous results ^9^. The participants with BD1 are plotted in their own group and together with BD, where they are the majority of the participants. MDD: Major depression disorder (n = 2,662) BD1: bipolar disorder type I (n = 2,298). BD: bipolar disorder (n = 1,742)., SCZ: Schizophrenia (n = 1,126), Other: includes schizoaffective disorder, delusional disorder and brief psychotic disorder (n= 657).

**Supplementary Figure 3. Comparison of prediction of diagnosis by SCZ, MDD, and BD1 polygenic scores in univariate and multivariate models.** Forest plot depicting odd ratios (OR) of the logistic regression of polygenic scores on MDD, SCZ, and BD1. Polygenic scores were calculated from external GWAS. The points represent the OR for each one standard deviation (SD) increase from the mean PRS of controls, and the error bars represent 95% confidence intervals. The vertical dashed line represents OR=1. PRS: polygenic risk score. BD1: bipolar disorder type I (light green), SCZ: schizophrenia (magenta), and MDD: major depression disorder (blue). Top row: univariate results, bottom row, multivariate results.

**Supplementary Figure 4. Correlations of individual PRS variance with genetic ancestry.** Individual variances were calculated for each PRS using PRScs posterior samples (n-iter=500, n-burnin=10,000) and correlated to global ancestry at the individual level. Y-axis is individual PRS variance and X-axis is global ancestry. Top: correlations of African ancestry with PRS_MDD_, Middle: correlations to European ancestry, Bottom: correlations to Native American ancestry.

**Supplementary Figure 5. Performance of ancestry-specific partial polygenic scores.** Measures of concordance and performance for partial polygenic scores. Partial polygenic scores were built from dosages of risk alleles on each respective ancestral background and using the weights of SMI GWAS. Only SNPs with information of local ancestry were used, and a total polygenic score was built by summing all the partial polygenic scores (PRS_TOTAL= pPRS_AFR_ + pPRS_EUR_ + pPRS_AMR_). A) Correlations between the traditional PRS calculated with PRScs and PRS_TOTAL. B) proportion of the contribution of partial PRS to the PRS_TOTAL. C) PRS were applied in a logistic regression with the respective diagnosis as outcome, previous regression of the first four PCs. Each plot (page) comprises the PRS of a diagnosis. MDD: Major depression disorder (BD1: bipolar disorder type I. BD: bipolar disorder type. SCZ: Schizophrenia.

**Supplementary Figure 6. Simulation of non-EUR partial polygenic scores in admixed genetic background.** Forest plot depicting log odds of diagnosis predictions with polygenic scores calculated with PRScs from the GWAS (PRScs), total polygenic scores which is the sum of the partial polygenic scores (PRS_TOTAL), partial non-European-specific polygenic scores (pPRS_AFR_, pPRS_AMR_), and the summarized 100 simulated non-EUR-specific polygenic scores (pPRS_AMR_* simulations in Supplementary Figure 7a, pPRS_AFR_* simulations in Supplementary Figure 7b). PRS were calculated from external GWAS, and the first four PCs were regressed out before application of logistic regression. Simulations were calculated by taking random sets of SNPs of the same size as the real non-EUR specific partial PRS and using their respective weights and EUR dosage to build PRS. For the simulations the point estimate represents the average log(OR) across simulations, and the error bar represents the first and third quartile of their distribution. The distribution of point estimates is represented in the gray density plot and the barcode below. For all other PRS error bars represent the 95% confidence interval.

**Supplementary Figure 7. Mean cognitive performance test scores by diagnosis.** Mean score of each cognitive performance test for each diagnosis (points) and confidence 95% confidence intervals (error bars). The scores were inverse normal transformed and standardized to the mean and standard deviation of controls. MDD: Major depression disorder, BD1: bipolar disorder type I, SCZ: Schizophrenia. ATT: Continuous performance test WM: Letter-N-back test. PS: Digit substitution test. FMEM: Face memory test. AM: Digit recall test, NVR: Matrix analysis test. EID: Emotion identification test. EDI: emotion differentiation test.

**Supplementary Figure 8a. Association of ancestry-specific partial polygenic scores of psychiatric diagnoses with transdiagnostic symptoms in cases.** Forest plots indicating odds ratio (OR, points) and 95% confidence intervals (error bars) of the logistic regression model of diagnosis ancestry-specific partial PRS on 14 transdiagnostic symptoms. All PRS were calculated from external GWAS, and the first four PCs, and diagnosis was regressed out before application of logistic regression. Partial PRS used the same process but using only the dosage (risk alleles) of the specific respective ancestry. Dashed lines indicate OR= 1.0. Associations under the multiple testing correction threshold (p < 3.23x10^-4^ for Admixed, P < 1.08X10^-4^ for specific ancestries) are indicated by the red asterisk, associations under the FDR<0.05 threshold with the blue asterisk. AnyHal: Any Hallucination, AnyDel: Any Delusion. EUR: European risk alleles only, AFR: African risk alleles only, AMR: Native American risk alleles only, Admixed: all the available risk alleles.

**Supplementary Figure 8b. Association of ancestry-specific partial polygenic scores of psychiatric diagnoses with CNB scores.** Forest plots indicating effect size (points) and 95% confidence intervals (error bars) of the logistic regression model of diagnosis ancestry-specific partial PRS on 17 speed and accuracy scores of the Penn Computerized Neurocognitive Battery (CNB). All PRS were calculated from external GWAS, and the first four PCs, and diagnosis was regressed out before application of linear regression. Partial PRS used the same process but using only the dosage (risk alleles) of the specific respective ancestry. Dashed lines indicate OR= 1.0. Associations under the multiple testing correction threshold (p < 3.23x10^-4^ for Admixed, P < 1.08X10^-4^ for specific ancestries) are indicated by the red asterisk, associations under the FDR<0.05 threshold with the blue asterisk. MatxAnalysis: Matrix analysis test. DigitRecall: Digit symbol test, recall trials. FaceMem: Face memory test. DigitSub: Digit symbol substitution test. LetterNB: Letter-N-back test. ContPerform: Continuous performance test. MotorPx: Motor praxis test. EmoID: Emotional identification test. EmoDiff: Measured emotion differentiation test. SD: Standard deviation. EUR: European risk alleles only, AFR: African risk alleles only, AMR: Native American risk alleles only, Admixed: all the available risk alleles.

**Supplementary Figure 9. Distribution of Higher Criticism across 35 traits.** The Higher Criticism statistics examines the departure of a set of p-values from the Uniform [0,1] (the distribution expected under the null of no association). Higher values represent stronger departures from the Uniform. Cognitive test abbreviations are as in Supplementary Figure 7.

**Supplementary Information**

*Detailed sample Ascertainment*

Cases with severe mental illness were ascertained through electronic health records (EHR) at Clínica San Juan de Dios de Manizales (CSJDM) in Manizales, Caldas and the Hospital Universitario San Vicente Fundación (HUSVF) in Medellín, Antioquía, beginning in 2017, and at Hospital Mental de Antioquia (HOMO) in Bello, Antioquia, beginning in 2019. Recruitment ended at all sites in February 2022. Individuals were invited to participate in the project based on the following criteria: (a) diagnosis of a severe mood or psychosis spectrum disorder, meeting at least one of the following five severity criteria (i) history of psychiatric hospitalization, (ii) treatment for symptoms warranting such admission, (iii) suicide attempt, (iv) presence of psychotic symptoms, and/or (v) history of electroconvulsive therapy; (b) presenting symptoms were not clearly caused by a substance use disorder, in the judgment of an evaluating clinician; (c) have two Paisa surnames ^19^ (d) aged 18 or above; (e) not a first-degree relative of another participant (f) understand and sign an informed consent document; (g) no intellectual disability, and (h) no history of serious brain trauma or neurological disorder.

Healthy controls were ascertained from the same communities as cases, and recruited from friends, neighbors, or in-laws of cases, or from university students/staff and hospital staff. All controls met the following criteria: (a) no (current or lifetime) severe mental illness, as evaluated through the overview screening module of the NetSCID (b) no current substance use disorder, and (c) fulfillment of criteria c-h described for cases. Cases and controls were reimbursed for transportation costs but were not otherwise compensated.
