## SupplementaryFigures for "Effect of ancestry and shared genetic architecture of serious mental illness on symptoms and cognition in an admixed Latin American population"

sFig 1.

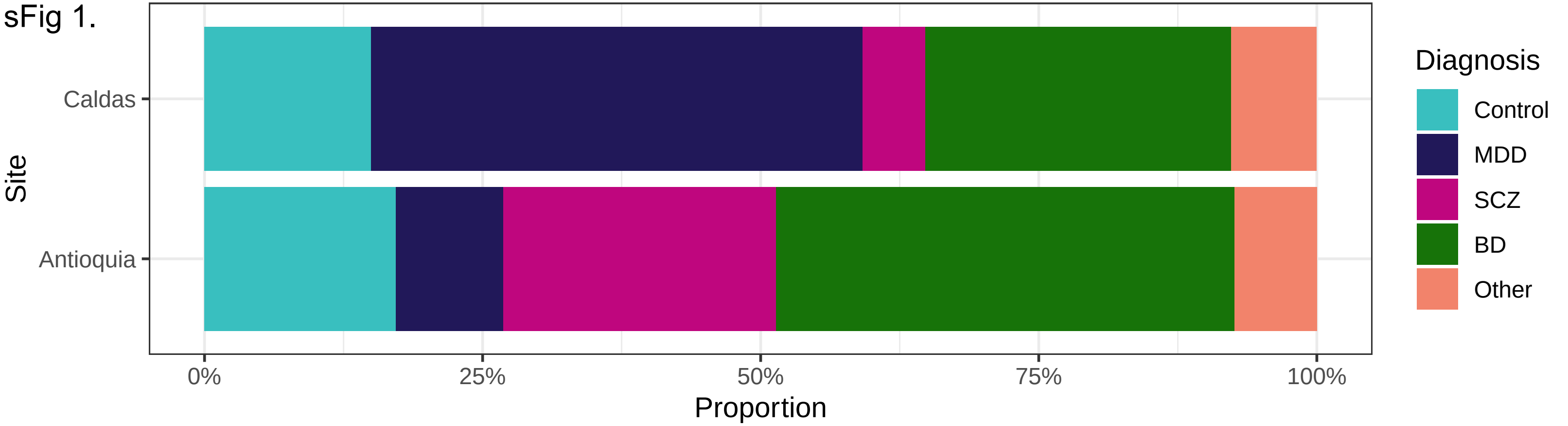
Diagnosis
Control
MDD
SCZ
BD
Other
Site
Caldas
Antioquia
0%
25%
50%
75%
100%
Proportion

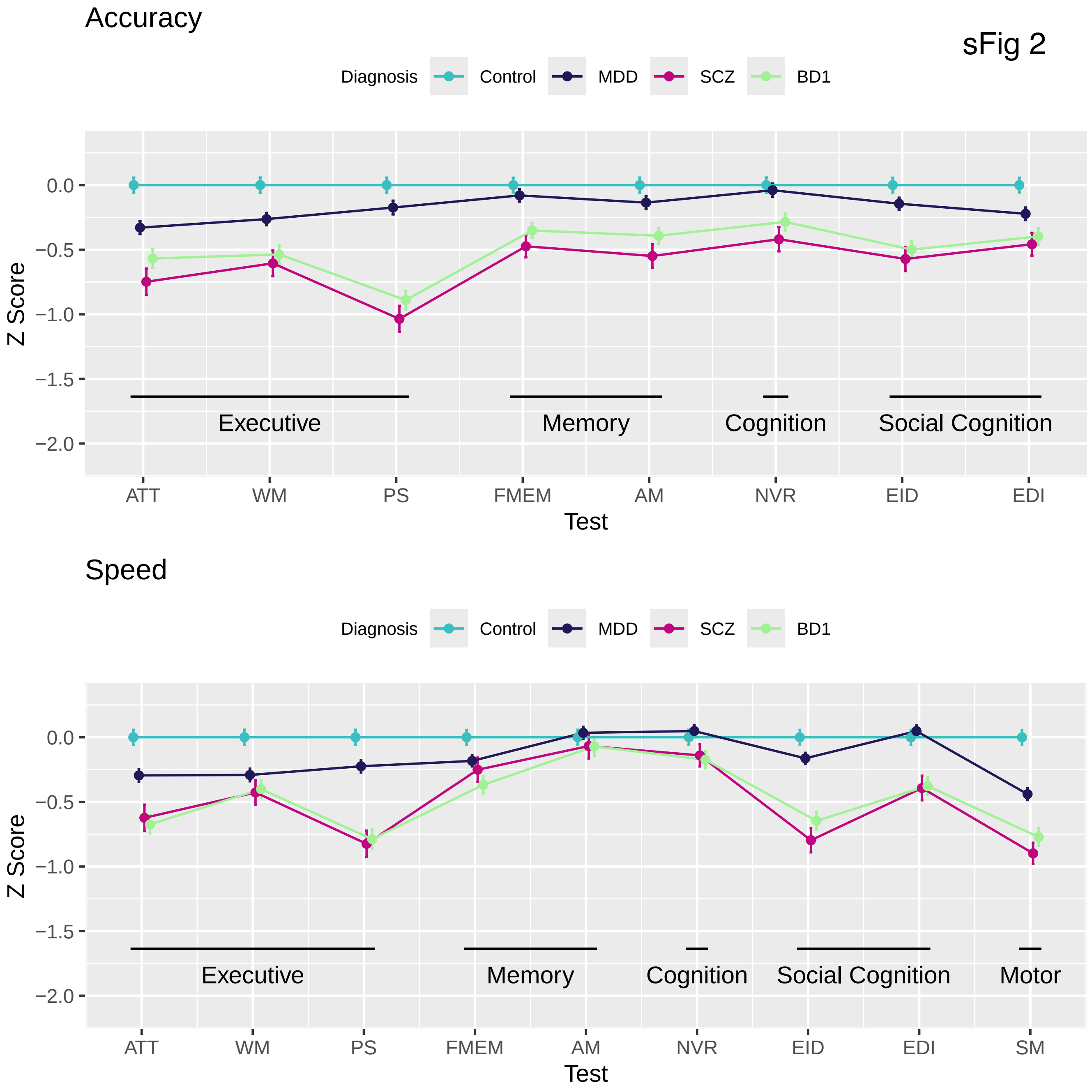
Accuracy
sFig 2
Diagnosis
Control
MDD
SCZ
BD1
0.0
−0.5
−1.0
−1.5
−2.0
Z Score
Executive
Memory
Cognition
Social Cognition
ATT
WM
PS
FMEM
AM
NVR
EID
EDI
Test
Speed
Diagnosis
Control
MDD
SCZ
BD1
0.0
−0.5
−1.0
−1.5
−2.0
Z Score
Executive
Memory
Cognition
Social Cognition
Motor
ATT
WM
PS
FMEM
AM
NVR
EID
EDI
SM
Test

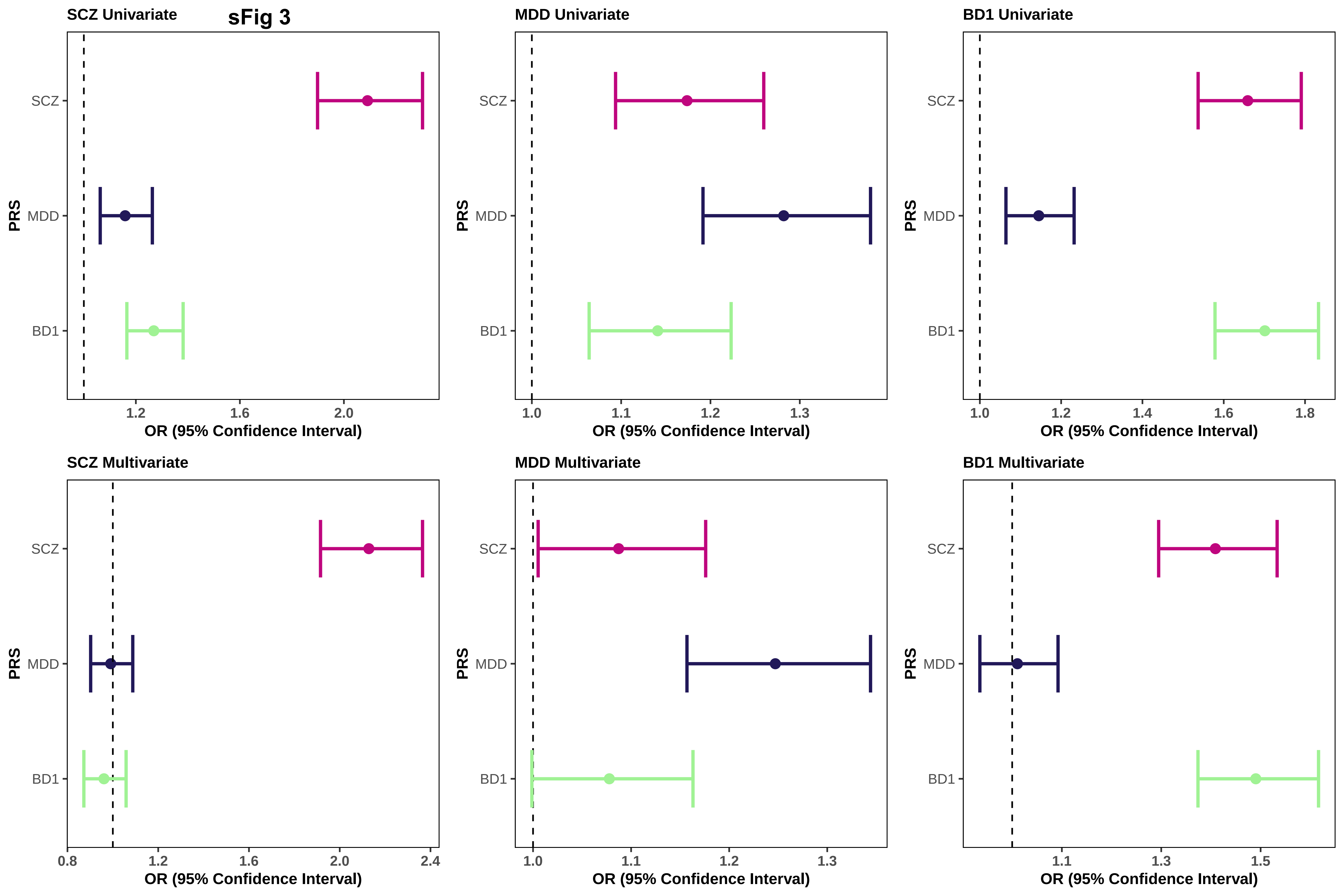
SCZ Univariate
sFig 3
MDD Univariate
BD1 Univariate
SCZ Multivariate
MDD Multivariate
BD1 Multivariate
PRS
SCZ
MDD
BD1
OR (95% Confidence Interval)
1.2
1.6
2.0
1.0
1.1
1.3
1.4
1.8
0.8
2.4
1.5

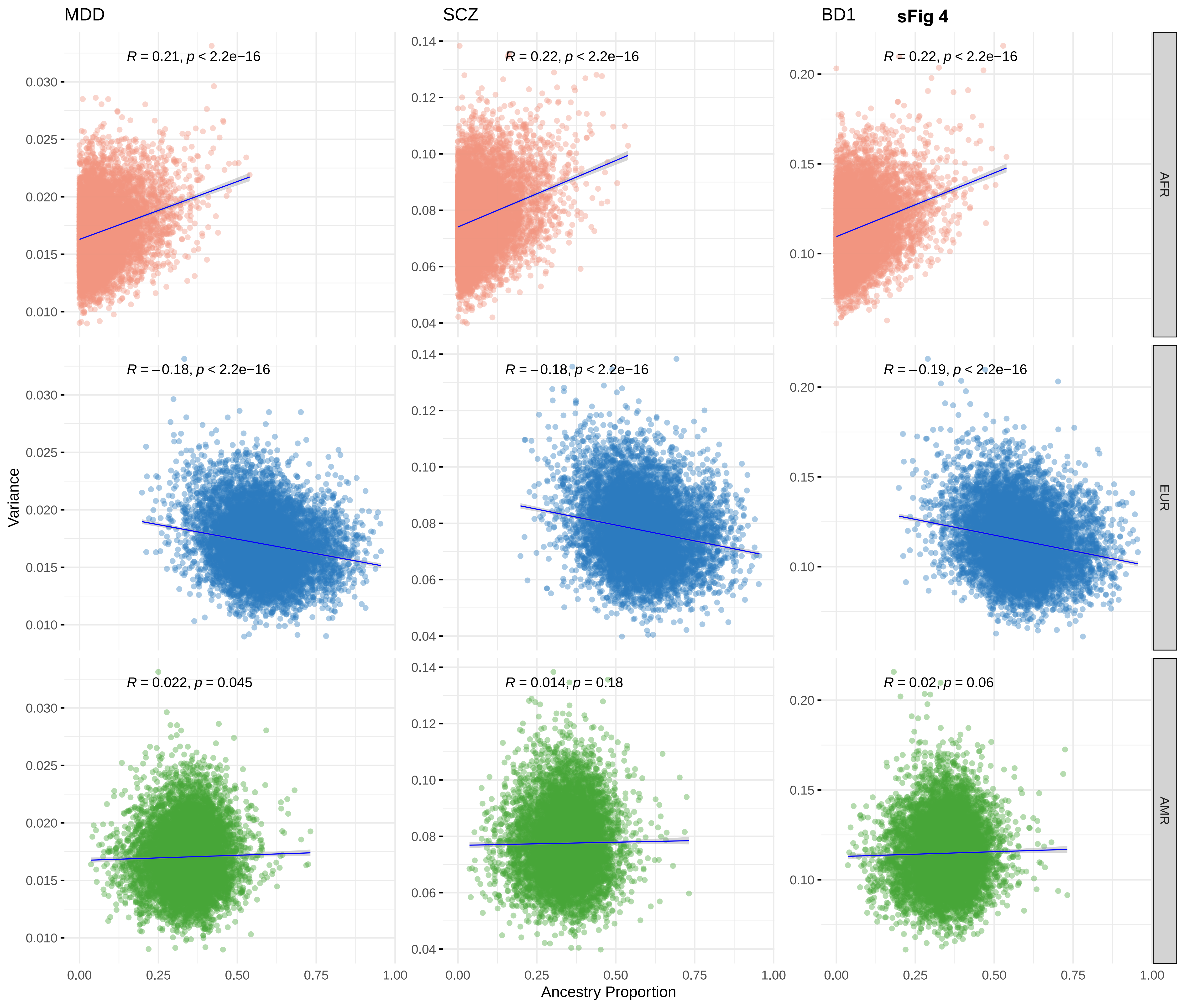

sFig 4
MDD
SCZ
BD1
AFR
EUR
AMR
R = 0.21, p < 2.2e−16
R = 0.22, p < 2.2e−16
R = 0.22, p < 2.2e−16
R = −0.18, p < 2.2e−16
R = −0.18, p < 2.2e−16
R = −0.19, p < 2.2e−16
R = 0.022, p = 0.045
R = 0.014, p = 0.18
R = 0.02, p = 0.06
Variance
Ancestry Proportion
0.00 0.25 0.50 0.75 1.00
0.010 0.015 0.020 0.025 0.030
0.04 0.06 0.08 0.10 0.12 0.14
0.10 0.15 0.20

**sFig 5.**

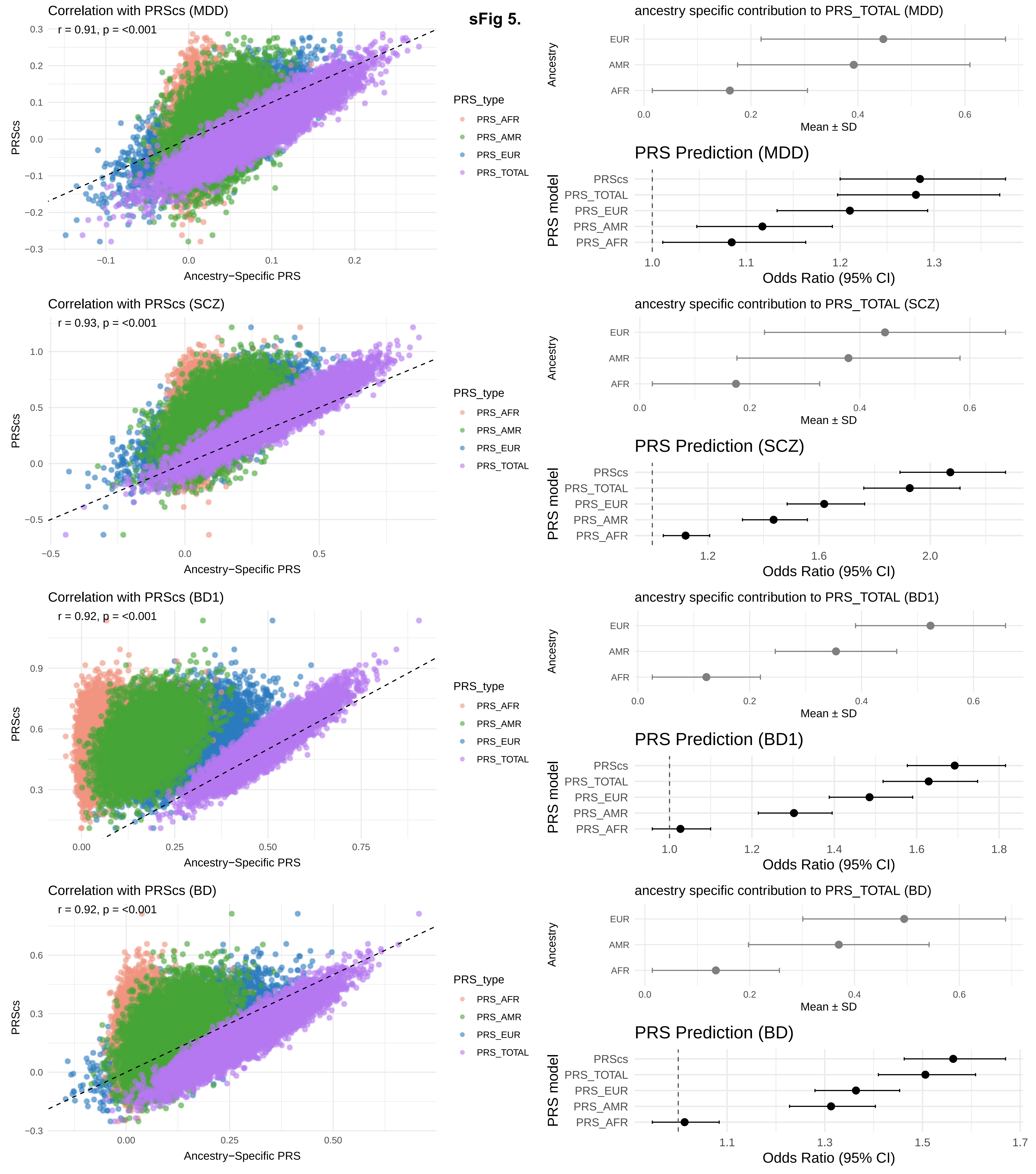

Correlation with PRScs (MDD)
r = 0.91, p = <0.001
PRScs
Ancestry−Specific PRS
PRS_type
PRS_AFR
PRS_AMR
PRS_EUR
PRS_TOTAL
ancestry specific contribution to PRS_TOTAL (MDD)
Ancestry
EUR
AMR
AFR
Mean ± SD
PRS Prediction (MDD)
PRS model
PRScs
PRS_TOTAL
PRS_EUR
PRS_AMR
PRS_AFR
Odds Ratio (95% CI)
Correlation with PRScs (SCZ)
r = 0.93, p = <0.001
ancestry specific contribution to PRS_TOTAL (SCZ)
PRS Prediction (SCZ)
Correlation with PRScs (BD1)
r = 0.92, p = <0.001
ancestry specific contribution to PRS_TOTAL (BD1)
PRS Prediction (BD1)
Correlation with PRScs (BD)
r = 0.92, p = <0.001
ancestry specific contribution to PRS_TOTAL (BD)
PRS Prediction (BD)

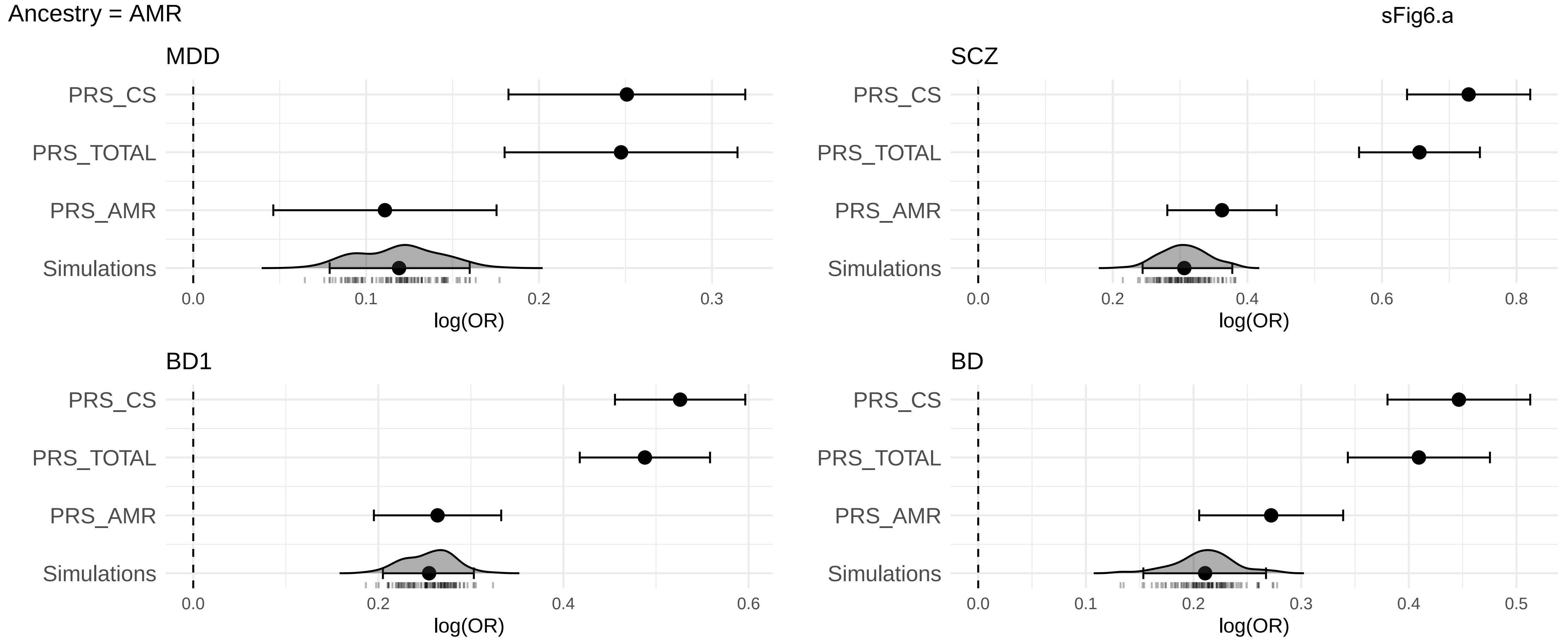

Ancestry = AMR
sFig6.a
MDD
SCZ
BD1
BD
PRS_CS
PRS_TOTAL
PRS_AMR
Simulations
log(OR)

Ancestry = AFR

sFig 6b.

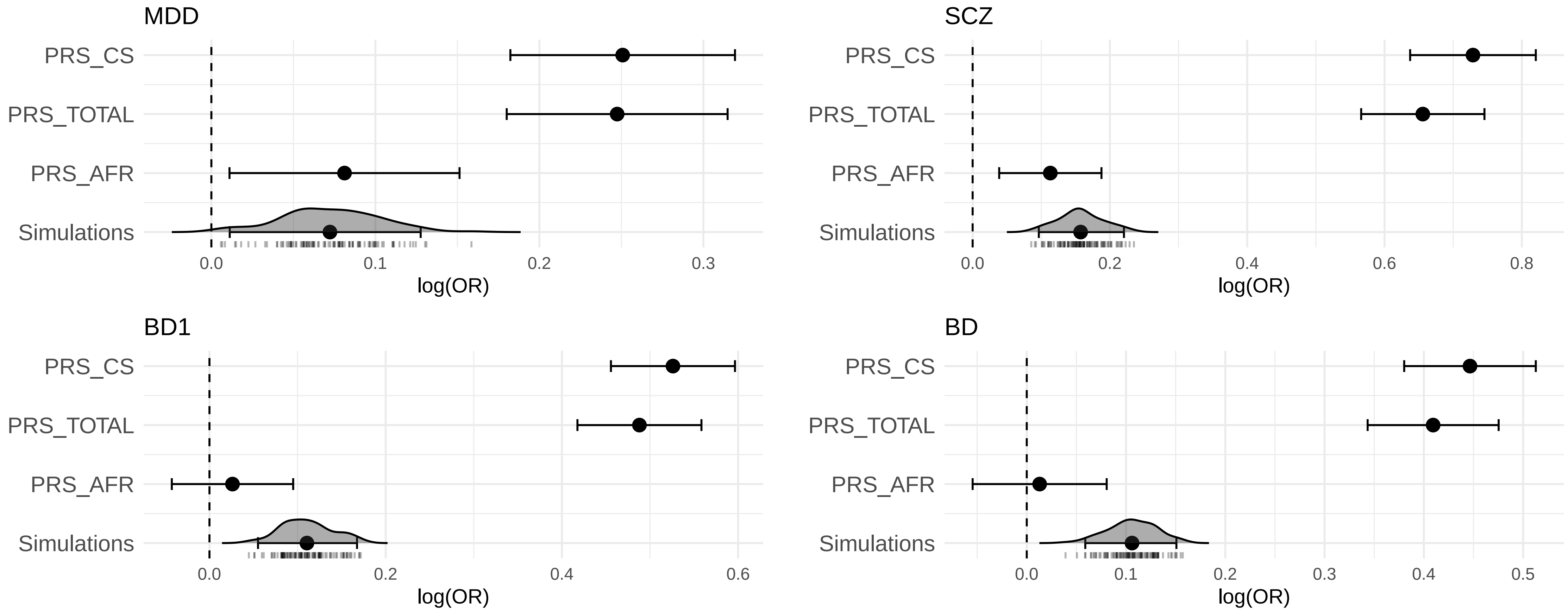

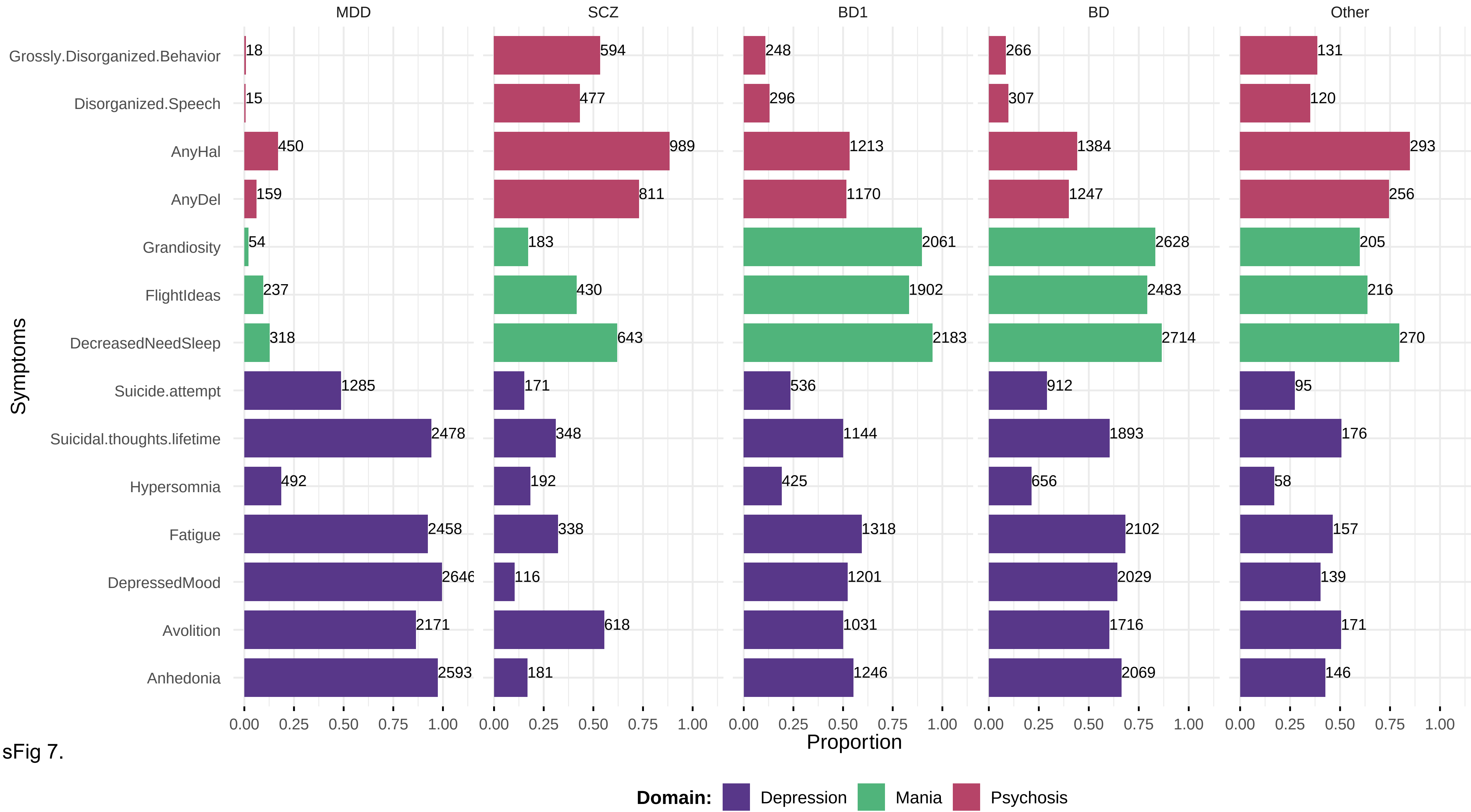

MDD
SCZ
BD1
BD
Other
Grossly.Disorganized.Behavior
Disorganized.Speech
AnyHal
AnyDel
Grandiosity
FlightIdeas
DecreasedNeedSleep
Suicide.attempt
Suicidal.thoughts.lifetime
Hypersomnia
Fatigue
DepressedMood
Avolition
Anhedonia
Symptoms
MDD: 18, 15, 450, 159, 54, 237, 318, 1285, 2478, 492, 2458, 2646, 2171, 2593
SCZ: 594, 477, 989, 811, 183, 430, 643, 171, 348, 192, 338, 116, 618, 181
BD1: 248, 296, 1213, 1170, 2061, 1902, 2183, 536, 1144, 425, 1318, 1201, 1031, 1246
BD: 266, 307, 1384, 1247, 2628, 2483, 2714, 912, 1893, 656, 2102, 2029, 1716, 2069
Other: 131, 120, 293, 256, 205, 216, 270, 95, 176, 58, 157, 139, 171, 146
0.00 0.25 0.50 0.75 1.00
Proportion
Domain: Depression Mania Psychosis


sFig 7.

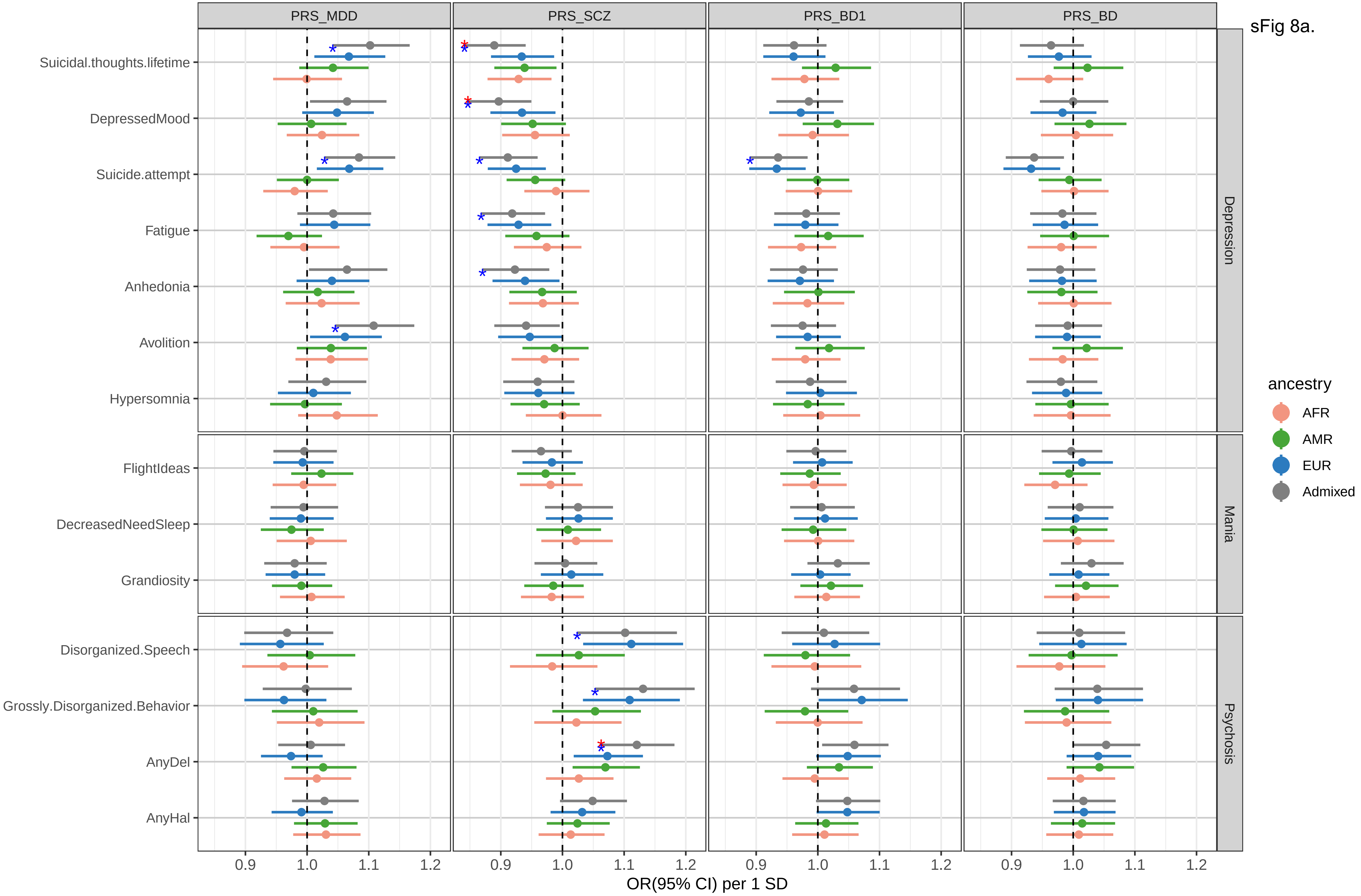
sFig 8a.
PRS_MDD
PRS_SCZ
PRS_BD1
PRS_BD
Suicidal.thoughts.lifetime
DepressedMood
Suicide.attempt
Fatigue
Anhedonia
Avolition
Hypersomnia
FlightIdeas
DecreasedNeedSleep
Grandiosity
Disorganized.Speech
Grossly.Disorganized.Behavior
AnyDel
AnyHal
Depression
Mania
Psychosis
ancestry
AFR
AMR
EUR
Admixed
0.9
1.0
1.1
1.2
OR(95% CI) per 1 SD

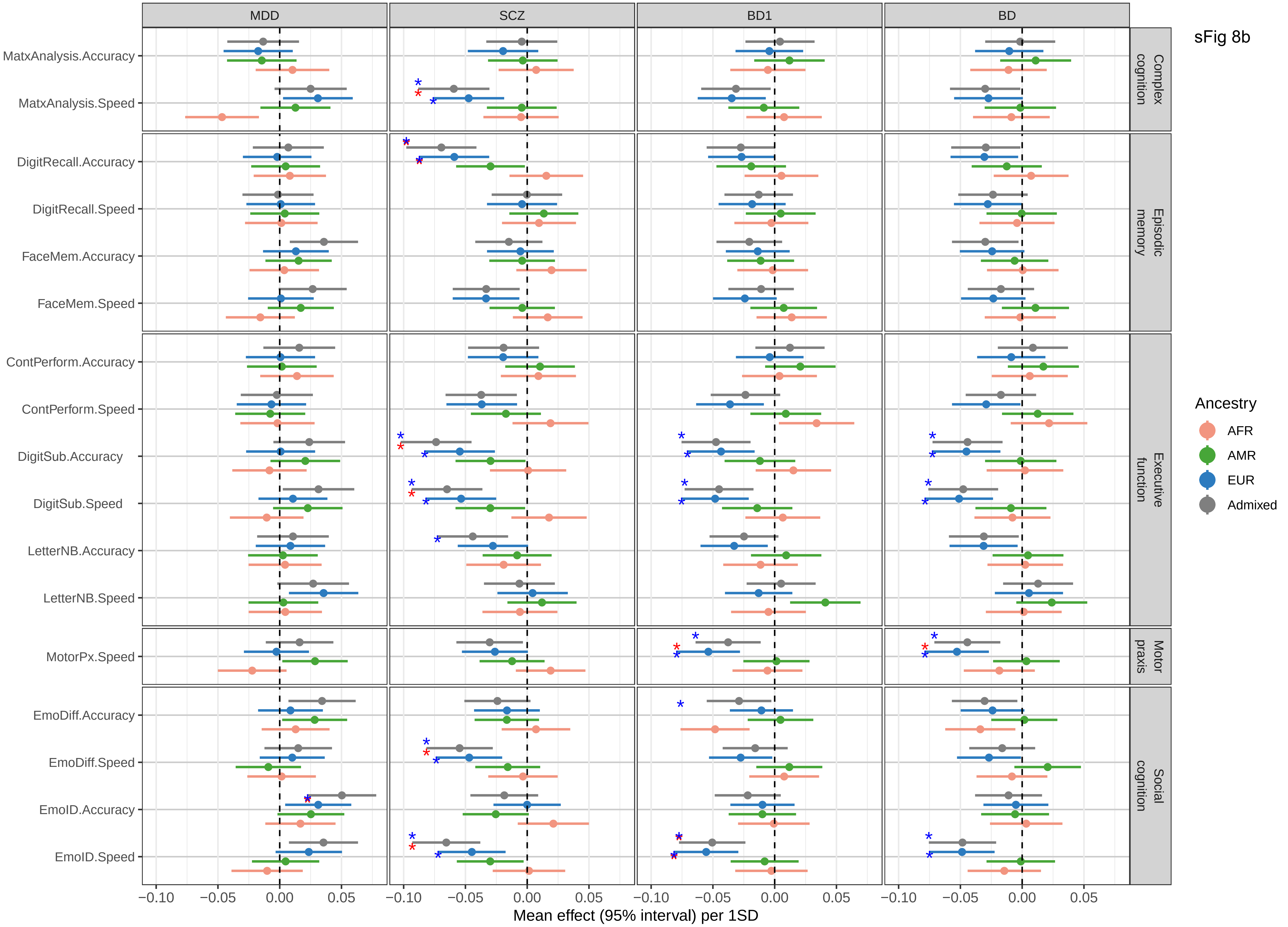

sFig 8b
MDD
SCZ
BD1
BD
Complex cognition
Episodic memory
Executive function
Motor praxis
Social cognition
MatxAnalysis.Accuracy
MatxAnalysis.Speed
DigitRecall.Accuracy
DigitRecall.Speed
FaceMem.Accuracy
FaceMem.Speed
ContPerform.Accuracy
ContPerform.Speed
DigitSub.Accuracy
DigitSub.Speed
LetterNB.Accuracy
LetterNB.Speed
MotorPx.Speed
EmoDiff.Accuracy
EmoDiff.Speed
EmoID.Accuracy
EmoID.Speed
−0.10
−0.05
0.00
0.05
Mean effect (95% interval) per 1SD
Ancestry
AFR
AMR
EUR
Admixed

sFig 9

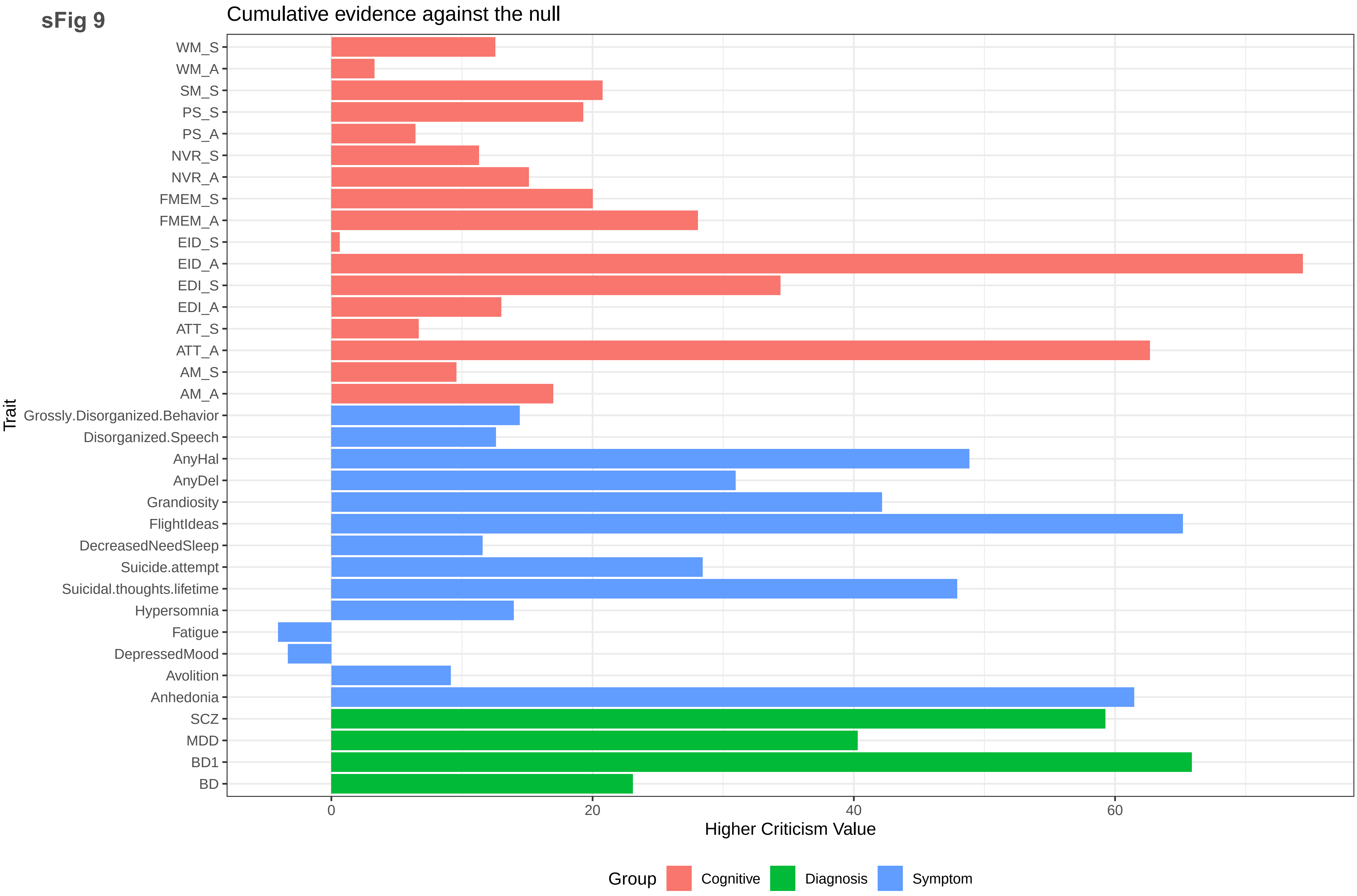
Cumulative evidence against the null
Trait
WM_S
WM_A
SM_S
PS_S
PS_A
NVR_S
NVR_A
FMEM_S
FMEM_A
EID_S
EID_A
EDI_S
EDI_A
ATT_S
ATT_A
AM_S
AM_A
Grossly.Disorganized.Behavior
Disorganized.Speech
AnyHal
AnyDel
Grandiosity
FlightIdeas
DecreasedNeedSleep
Suicide.attempt
Suicidal.thoughts.lifetime
Hypersomnia
Fatigue
DepressedMood
Avolition
Anhedonia
SCZ
MDD
BD1
BD
0
20
40
60
Higher Criticism Value
Group
Cognitive
Diagnosis
Symptom
